# Assessing the transportability of clinical prediction models for cognitive impairment using causal models

**DOI:** 10.1101/2022.03.01.22271617

**Authors:** Jana Fehr, Marco Piccininni, Tobias Kurth, Stefan Konigorski, the Alzheimer’s Disease Neuroimaging Initiative

## Abstract

**Background:** Machine learning models promise to support diagnostic predictions, but may not perform well in new settings. Selecting the best model for a new setting without available data is challenging. We aimed to investigate the transportability by calibration and discrimination of prediction models for cognitive impairment in simulated external settings with different distributions of demographic and clinical characteristics.

**Methods:** We mapped and quantified relationships between variables associated with cognitive impairment using causal graphs, structural equation models, and data from the ADNI study. These estimates were then used to generate datasets and evaluate prediction models with different sets of predictors. We measured transportability to external settings under guided interventions on age, APOE ε4, and tau-protein, using differences between internal and external settings measured by calibration metrics and area under the receiver operating curve (AUC).

**Results:** Calibration differences indicated that models predicting with causes of the outcome were more transportable than those predicting with consequences. AUC differences indicated inconsistent trends of transportability between the different external settings. Models predicting with consequences tended to show higher AUC in the external settings compared to internal settings, while models with parents or all variables showed similar AUC.

**Conclusions:** We demonstrated with a practical prediction task example that predicting with causes of the outcome results in better transportability compared to anti-causal predictions measured by calibration differences. We conclude that calibration differences are more suitable than AUC differences to assess model transportability to external settings.

## Introduction

Dementia is the second leading cause of death globally,^1^ and more than 55 million people currently have dementia. Detecting dementia at an early stage of cognitive impairment is essential to give affected individuals adequate care and eventually administer disease-modifying treatments.^2^ In recent years, several machine learning (ML) models have been proposed to support clinical decision making by predicting the diagnosis of Alzheimer’s disease (AD) and cognitive impairment.^3–8^ The models were developed with data from different cohorts and included different predictor variables, such as image-derived brain volumetric measures, cognitive test results, or demographic predictors. One obstacle for deploying such prediction models in clinical practice is that they might not generalize well when being transported (i.e., being applied) to other settings (e.g., in another hospital or regions with different patient demographics). One reason for reduced transportability may be that ML models learn non-causal associations between input and output variables, which might be different in external settings.^9,10^ This scenario can especially occur when models predict a diagnosis based on clinical consequences of the disease (e.g., when prediction is in the anti-causal direction).^11–13^ For prospective applications, end-users face the challenge of finding the most transportable model to their setting where data has not yet been collected.

Causality research established two approaches to improve transportability for prediction models. First, causal relationships can be incorporated in prediction models *a priori* for learning relationships that are more stable across settings and can therefore avoid systematic failures in external settings.^10,14–17^ To this aim, directed acyclic graphs (DAGs) are a useful tool to map assumed causal relationships between variables, represent differences and commonalities between settings,^18,19^ and select variables for transportable health prediction tasks.^20–22^ Second, the causal validity of learned relationships can be assessed through guided interventions (also known as perturbations) on data distributions to simulate differences between internal and external validation settings.^17,23,24^ Research in epidemiology and ML has adopted DAGs and interventions to develop transportable ML models.^25–34^ Piccininni et al. described the use of DAGs for selecting a single predictor in a hypothetical clinical risk prediction model for AD.^25^ They discussed that prediction models for AD are more likely to transport well to different settings when the selected predictor is a cause of AD and not a consequence. Rojas-Carulla et al. and Magliacane et al. applied automatic hypothesis testing to determine a transportable predictor set across multiple source domains.^30,31^ Subbaswamy et al. used interventions to achieve that predictors do not depend on unreliable parts of the data-generating process and thereby generalize to unknown test data.^35^ In another study, Singh et al. proposed a model predicting acute kidney injury to ensure the fairness of predictions on unseen test data by applying DAGs and interventions.^32^ Steingrimsson et al. demonstrated an approach to assess transportability to external settings where the outcome variable has not yet been measured using inverse-odds weights.^36^ These and other works^37–39^ measured transportability to unseen data by comparing mean-squared error or discrimination error (e.g., area under the receiver operating curve (AUC)) between internal and external validation settings. Following the work of Van Calster et al., however, we argue that discriminatory metrics may not be a suitable metric to assess external validity because predicted risks can be unreliable even if algorithms have good discrimination, and instead suggest calibration as a suitable metric to measure transportability.^40^

In this work, we aim to compare the transportability, measured by calibration and discrimination, of models predicting cognitive impairment with different sets of predictors to simulated external settings to add evidence for two questions: 1) how to construct transportable ML models and 2) how to assess the transportability.

## Methods

### Data source and data preprocessing

Data used in the preparation of this article were obtained from the Alzheimer’s Disease Neuroimaging Initiative (ADNI) database (adni.loni.usc.edu). The primary goal of ADNI has been to test whether medical imaging, biological markers, and clinical and neuropsychological assessment can be combined to measure the progression of mild cognitive impairment (MCI) and early AD. For up-to-date information, see www.adni-info.org. The ADNI study acquired multiple longitudinal measurements from elderly subjects across more than 50 clinics in USA and Canada.^41,42^ The ADNI study enrolled participants who have mild cognitive impairment or a diagnosis of early AD and elderly controls between the age of 55 and 90 years old. We used the ADNI data subset that was created for the TADPOLE grand challenge (https://tadpole.grand-challenge.org/Data/), which is also available at https://ida.loni.usc.edu. We selected individuals who had a diagnosis and clinical measurements at baseline (n=1,737) and considered baseline variables that have been reported to be related to AD and had less than 30% of missing entries. Detailed information on data preprocessing is provided in Supplementary Text 1. Missing data were imputed using the R package ‘mice’ with default settings, and three imputed datasets were generated. All numeric variables were normalized by z-transformation. We defined the outcome variable ‘cognitive impairment’ based on the final diagnosis variable in ADNI by considering participants with cognitive normal or subjective memory complaint as “cognitively normal” and participants with mild cognitive impairment or AD as “with cognitive impairment.”

### DAG creation

In DAGs, nodes represent variables, and directed edges represent direct causal relationships pointing from the direct cause to the effect.^18,43,44^ We reviewed the scientific literature to identify causal relationships between variables in our dataset that are involved in cognitive impairment and AD processes (Supplementary Table S1) and mapped them in a first DAG (Supplementary Figure S1). Then, we tested if the generated DAG was a good fit to the ADNI dataset, using conditional independence testing with the R package ‘dagitty.’^45^ We reviewed those test results with low p-values and large point estimates, which indicated a violation of the implied conditional independence. We added 13 causal connections (Supplementary Text 2) according to the test results and our domain expertise to create the final DAG (Figure 1).

**Figure 1:**
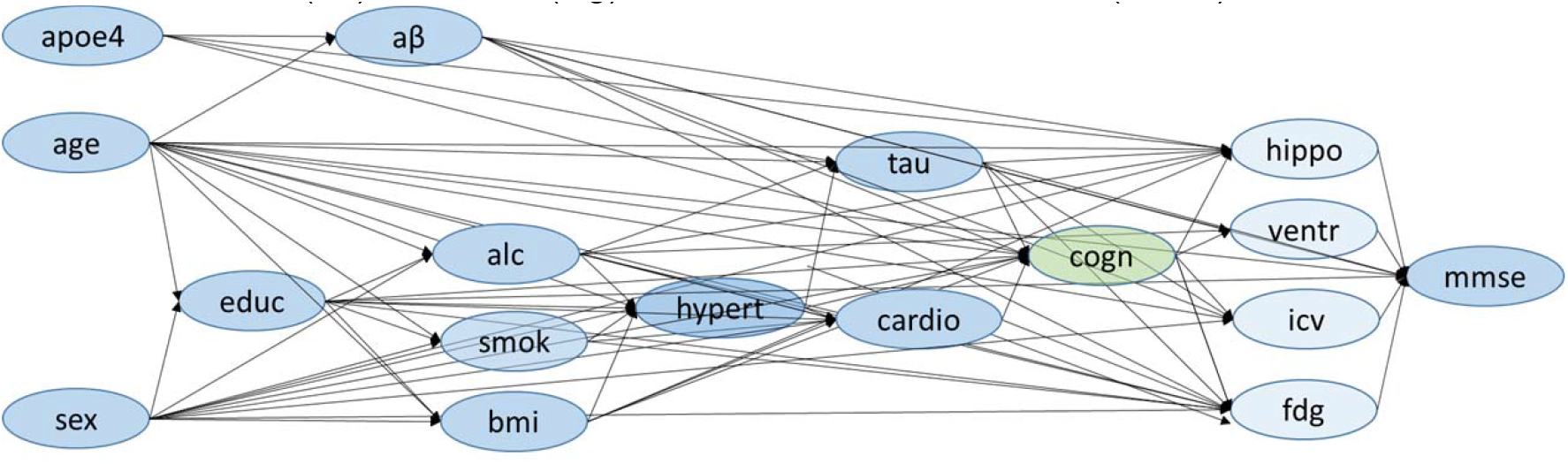
Directed acyclic graph of variables related to cognitive status. Predictor variables are marked in blue, and the outcome variable (cognitive status) in green. Directed arrows indicate assumed causal relationships between variables. The included variables are: APOE ε4 (apoe4), age, sex, education (educ), CSF-Aβ (aβ), history of alcohol abuse (alc), history of smoking behavior (smok), Body Mass Index (bmi), history of hypertension (hypert), CSF-tau (tau), history of cardiovascular events (cardio), cognitive status (cogn), hippocampus (hippo), ventricles (ventr), intracranial volume (icv), FDG-PET (fdg), Mini-Mental State Exam score (mmse).

### Semi-synthetic data generation using structural equation models

We fitted a structural equation model (SEM) using the three imputed ADNI datasets to quantify the causal relationships specified in our DAG. The SEM was implemented using the ‘sem’ function in the R package ‘lavaan’ with default parameters.^46^ For numeric endogenous (dependent) variables, the function computes weighted least squares estimates. For categorical endogenous variables, the function automatically uses a diagonally weighted least squares estimator and assumes that a conditional normally distributed latent variable underlies the categorical variable (and estimates the thresholds).

We then used the SEM parameter estimates (Supplementary Table S2) to generate six semi-synthetic datasets with 10,000 individuals each: one for training, one for internal validation, and four for external validation of ML models. We bootstrapped exogenous (independent) variables (age, sex, and APOE ε4) 10,000 times without replacement from the original data. We used those to generate the endogenous variables for training and internal validation sets, using the linear equations from the SEM (Supplementary Figure S2). We generate the four external validation sets implemented by interventions on the variables to reflect different populations with

1. a younger mean age, compared to the original data (73 years ⇒ 35 years, which is similar to the global world population mean at 31^1^),
2. a younger mean age compared to original data, but higher compared to the first age validation setting ⇒ 65 years (referred to as “age2”),
3. lower prevalence of the APOE ε4 gene compared to the original data (46.9% ⇒ 5.0%), and
4. a different mechanism generating the endogenous variable tau-protein, measured in cerebrospinal fluid.

For the external age-intervention data, we sampled the age variable from a normal distribution with a mean age of 35 (and mean of 65 for age2 setting) and standard deviation of 10 and bootstrapped APOE ε4 and sex. For the APOE ε4 intervention, we sampled from a Bernoulli distribution with a 5% probability. For the tau intervention, we altered the mechanism determining tau levels by intervening on the parameters estimated by the SEM for the tau equation. In particular, we arbitrarily changed the intercept from the tau equation from -0.5 to 0.9, increased the influence of age on tau from 0.37 to 0.9, increased the influence of apoe4 from 0.57 to 0.8, increased the influence of hypertension from 0.14 to 0.9, and reduced the influence of alcohol from 0.57 to 0.001.

### Prediction algorithms

We applied logistic regression, lasso regression, random forest, and generalized boosted regression (GBM) to predict the cognitive state of an individual as either cognitive normal or with cognitive impairment. Logistic regression was performed using the glm function in the ‘stats’ R package. Lasso regression was implemented using the ‘glmnet’ R package.^47^ The lasso model was initialized with an optimized penalization hyperparameter obtained from a grid-search with 10-fold cross validation that selected the value of lambda for minimum deviance. The random forest is an ensemble of regression trees, which aims at improving generalizability compared to a single regression tree.^48^ Previous works demonstrated the strengths of random forests for diagnostic prediction modelling of AD.^3–5^ The random forest algorithm was applied from the ‘randomForest’ R package, using 500 trees and v*p* number of variables randomly sampled at each split (as per default), where *p* is the number of predictors. GBM implements boosting by adding regression trees sequentially with respect to the error of the current tree ensemble. This boosting approach increases robustness and generalizability compared to a single regression tree.^49–51^ The GBM algorithm was applied using the ‘gbm’ R package with 100 trees (as per default).

Based on the causal assumptions in our DAG, we defined four predictor sets that included either all variables or only those which are direct causes of the outcome (defined as parent nodes), or only direct consequences of the outcome (defined as children nodes), or only exogenous variables (age, sex, and APOE ε4) (Table 1). Each ML model was trained and validated with each predictor set. We performed 10,000 repetitions to generate the six datasets (one for training, one for internal validation, and four for external validation) for training and validating all prediction models.

**Table 1:** Predictor sets with corresponding lists of variable names. The predictor variables comprised age, APOE ε4, sex, years of education, Body Mass Index (BMI), history of hypertension, history of alcohol abuse, history of smoking behavior, history of cardiovascular events, Cerebrospinal fluid (CSF) of tau and Amyloid β (Aβ), volumes measured from brain magnetic resonance imaging (sum of left and right hippocampal volumes, sum of left and right ventricle volumes, intracranial volume, fluorodeoxy-glucose-positron emission tomography (FDG-PET)), and Mini Mental State Exam score (MMSE).

| Predictor set | Variable names |
| --- | --- |
| All nodes | age, APOE $\epsilon 4$, sex, education, BMI, history of hypertension, history of alcohol abuse, history of smoking behavior, history of cardiovascular events, CSF-tau, CSF-$A\beta$, hippocampus, ventricles, intracranial volume, FDG-PET, MMSE |
| Parent nodes | age, APOE $\epsilon 4$, sex, education, BMI, history of cardiovascular events, CSF-tau, CSF-$A\beta$ |
| Children nodes | hippocampus, ventricles, intracranial volume, FDG-PET, MMSE |
| Exogenous nodes | age, APOE $\epsilon 4$, sex |

We repeated this procedure three times, each time using the SEM parameters obtained from one of the three imputed ADNI datasets. As a sensitivity analysis, we additionally ran 100 repetitions using hyperparameter tuning to minimize the deviance for the random forest (tuned parameters: number of predictors sampled for splitting at each node from 1 to 5, and the minimum size of terminal nodes of 1, 5 or 10) using generated training datasets with 1,000 observations.

### Assessing transportability

We calculated calibration metrics and the AUC discrimination performance for all prediction models in the internal validation setting and in each external validation setting. Calibration was measured using the Integrated Calibration Index (ICI)^52^ and the calibration component of a three-way decomposed Brier score.^53^ Low ICI and Brier scores indicate better calibration. The Brier calibration component (termed ‘reliability’) was obtained from the bias-corrected ‘BrierDecomp’ function of the ‘SpecsVerification’ R package using quantile bins of predicted probabilities in 10% steps. The AUC was obtained from the ‘pROC’ R package. AUC values close to one indicate good discrimination performance.

We assessed the transportability between the internal setting and each external validation setting by differences in calibration (ICI or Brier score) and in AUC. Differences of zero indicate equal performance in both internal validation and external settings and, therefore, good transportability. Negative calibration differences values indicate decreased calibration from internal validation to the intervention setting and therefore decreased transportability. AUC differences greater than zero indicate decreased discriminatory performance from internal validation to the intervention setting and thus low transportability.

We calculated the median, 2.5%, and 97.5% percentiles for performance metrics across all 10,000 repetitions for each of the three imputed ADNI datasets. See Figure 2 for a summary of our workflow. All prediction algorithms, data simulations, and data analyses were implemented using R version 4.0.3.

**Figure 2:**
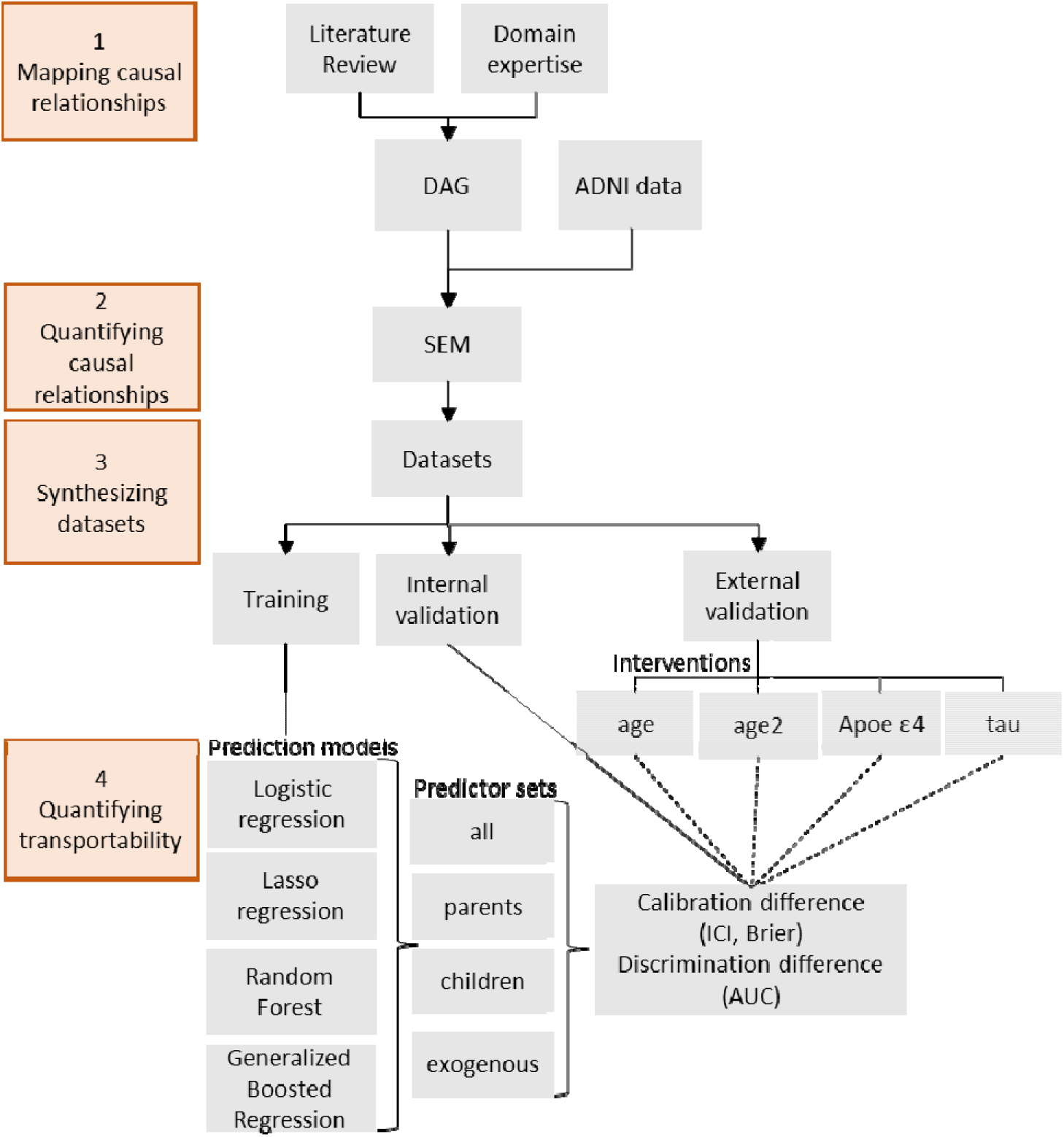
Our approach to assess the transportability of machine learning models predicting cognitive impairment. Orange boxes mark the four general steps of our workflow. We first mapped knowledge about cognitive impairment into a Directed Acyclic Graph (DAG) and quantified those using Structural equation modelling (SEM) and data from the Alzheimer’s Disease Neuroimaging Initiative (ADNI). The estimates were used in linear equations to generate datasets for training, internal validation, and four external validation datasets with interventions on age, APOE ε4, and tau. The age variable was intervened two times (age and age2) by sampling from normal distributions with two different mean age values (35 and 65). We trained four machine learning algorithms (logistic regression, lasso regression, random forest, and generalized boosted regression) to predict cognitive impairment using four sets of predictors. We measured transportability between internal and external settings using calibration differences, measured by Integrated Calibration Index (ICI) and Brier score, and differences in Area under the Receiver Operating Curve (AUC). Steps 3 to 4 (data synthesis and model training and validation) were repeated 10,000 times for each of the three imputed ADNI datasets.

## Results

### Description of the participants’ characteristics

The ADNI study, represented in TADPOLE, recorded a total of 1737 participants with a diagnosis at baseline together with their demographic information (age, sex, and education), behavioral information (smoking and alcohol abuse history), clinical measurements (BMI, FDG-PET imaging, brain volumetric measurements with MR imaging, Aβ and tau protein concentrations in cerebrospinal fluid (CSF), Minimental State Cognitive Exam (MMSE) and medical history (history of hypertension and cardiovascular events) (Table 2). Among all participants, 1214 (69.9%) were diagnosed with cognitive impairment, which comprised 872 (71.8%) individuals with mild cognitive impairment and 342 (28.7%) individuals with AD. A total of 523 (30.1%) individuals was diagnosed as cognitive normal, including 106 (20.3%) individuals with subjective memory complaint.

**Table 2:** Participant characteristics of ADNI dataset at baseline stratified by cognitive status. Numeric variables are indicated with * and are given with median and 25-75% interquartile range (IQR). All other variables are categorical variables with two categories, and the absolute number and the column-wise percentage of the reference category are given. Absolute numbers of missing values (Nmiss) are given. Abbreviations: Body Mass Index (BMI), Cerebrospinal fluid (CSF), Amyloid β (Aβ), Mini Mental State Exam score (MMSE).

| Variables | Cognitive normal<br>n=523 (30.1%) | Cognitive impairment<br>n=1214 (69.9%) | Total<br>n=1737 (100%) |
| --- | --- | --- | --- |
| Age* | 73.7 (70.5, 78.0) | 74.0 (68.3, 79.3) | 73.9 (69.2, 78.9) |
| APOE ε4 | 149 (28.6%)<br>Nmiss: 2 | 660 (54.8%)<br>Nmiss: 10 | 809 (46.9%)<br>Nmiss:12 |
| sex (male) | 253 (48.4%) | 704 (58.0%) | 957 (55.1%) |
| education* (years) | 16 (14, 18) | 16 (14, 18) | 16 (14, 18) |
| CSF-Aβ* (pg/ml) | 1271 (820.8, 1734.0)<br>Nmiss: 156 | 741.5 (559.1, 1130.3)<br>Nmiss: 366 | 854.2 (596.2, 1395.5)<br>Nmiss: 522 |
| CSF-tau* (pg/ml) | 214.3 (175.2, 287.8)<br>Nmiss: 156 | 281.6 (210.4, 379.2)<br>Nmiss: 366 | 257.8 (193.4, 349.7)<br>Nmiss: 522 |
| history of alcohol abuse | 16 (3.1%)<br>Nmiss: 6 | 27 (2.3%)<br>Nmiss: 27 | 43 (2.5%)<br>Nmiss: 33 |
| history of smoking | 115 (26.8%)<br>Nmiss: 94 | 223 (23.2%)<br>Nmiss: 254 | 338 (24.3%)<br>Nmiss: 348 |
| BMI* | 28.6 (25.8, 32.5)<br>Nmiss: 4 | 28.17 (25.5, 31.2)<br>Nmiss: 5 | 28.31 (25.5, 31.6)<br>Nmiss: 9 |
| history of hypertension | 130 (24.9%) | 460 (38.0%)<br>Nmiss: 5 | 590 (34.1%)<br>Nmiss: 5 |
| history of cardiovascular events | 343 (65.6%) | 835 (68.8%) | 1178 (67.8%) |
| MMSE* | 29.0 (29.0, 30.0) | 27.0 (25.0, 29.0) | 28.0 (25.0, 29.0) |

### Semi-synthetic data generation

The SEM estimated all parameters quantifying the causal relationships in the DAG in Figure 1. We reviewed the estimated parameters and found that many were in agreement with existing neurological domain knowledge. For example, age had a positive coefficient and therefore increased CSF-tau (0.37), the likelihood of hypertension (0.11), and the likelihood of cardiovascular events history (0.13) (Supplementary Table S3). Some estimated relationships, however, were controversial to domain knowledge. For example, increasing age decreased CSF-Aβ (−0.14) and the likelihood of cognitive impairment (−0.13). The SEM additionally indicated a small correlation between sex and age and between age and APOE ε4.

We compared endogenous variable distributions between the original ADNI data and generated validation datasets generated by SEM parameters from the first imputed dataset (Supplementary Table S4) and found that the percentage of cognitive impairment was similar between the internal validation set (70.0%, 2.5% and 97.5% percentiles [69.1, 71.0]) and the original ADNI data (69.9%). We further compared endogenous variable distributions between internal and external datasets. Lowering the mean age of 73.8 years in the internal validation setting to 35 years in the external setting slightly decreased the prevalence of cognitive impairment from 70.0% to 68.3%, increased the smoking prevalence from 23.4% to 34.1% and alcohol abuse history from 1.5% to 46.5%, decreased the prevalence of hypertension from 35.0% to 8.0% and previous cardiovascular events from 65.1% to 32.4%. Intervening on age increased the mean of Aβ from 1076.6 to 1521.0 pg/ml, shrank the mean of tau from 292.7 to 180.8 pg/ml, and increased the MMSE from 27.3 to 29.6, in comparison to the internal validation data. The differences between the internal and external variable distributions were smaller but followed the same trends when reducing the mean population age only slightly from 73.8 to 65.0 in the age2 intervention setting.

In the APOE ε4 intervention, lowering the prevalence of the APOE ε4 gene from the internal setting to the external setting decreased the prevalence of cognitive impairment from 70.0% to 63.1%, increased the mean of CSF-Aβ from 1076.6 to 1291.2 pg/ml and decreased the mean CSF-tau from 292.7 to 260.1 pg/ml.

In the tau intervention, altering the intercept and coefficients that determine tau levels between the internal setting to the external setting increased the level of CSF-tau from 292.7 to 476.4. Also they increased the prevalence of cognitive impairment from 70.0% to 82.3%.

### Internal calibration and discrimination of the models

We evaluated the internal validation performance measured by calibration and AUC metrics (Figure 3, Supplementary Figure S3, Supplementary Figure S4, and Supplementary Table S5) of all models. ICI scores were close to 0 for all logistic and lasso models (e.g., logistic and lasso with parents as predictors 0.009 2.5% and 97.5% percentiles [0.004, 0.016]). Random forest and GBM models had lower calibration (indicated by higher ICI values) compared to logistic and lasso models (using parents as predictors, random forest 0.035 [0.024, 0.046], GBM 0.029 [0.019, 0.039]). Random forest models predicting with exogenous variables (age, sex, and APOE ε4) had very low calibration (0.292 [0.277, 0.305]). Brier scores were exactly zero or closer to zero in the internal validation setting compared to the ICI scores, except for random forest models predicting with exogenous variables, which showed the lowest calibration (0.088 [0.080, 0.094]) among all models. AUC indicated the best performance for models predicting with all variables, and of those models, logistic regression and lasso regression achieved the highest AUC (0.75 [0.73, 0.76]) compared to random forest (0.73 [0.72, 0.74]) and GBM (0.71 [0.70, 0.73]). All models predicting with exogenous variables had the lowest discriminatory performance of 0.5.

**Figure 3:**
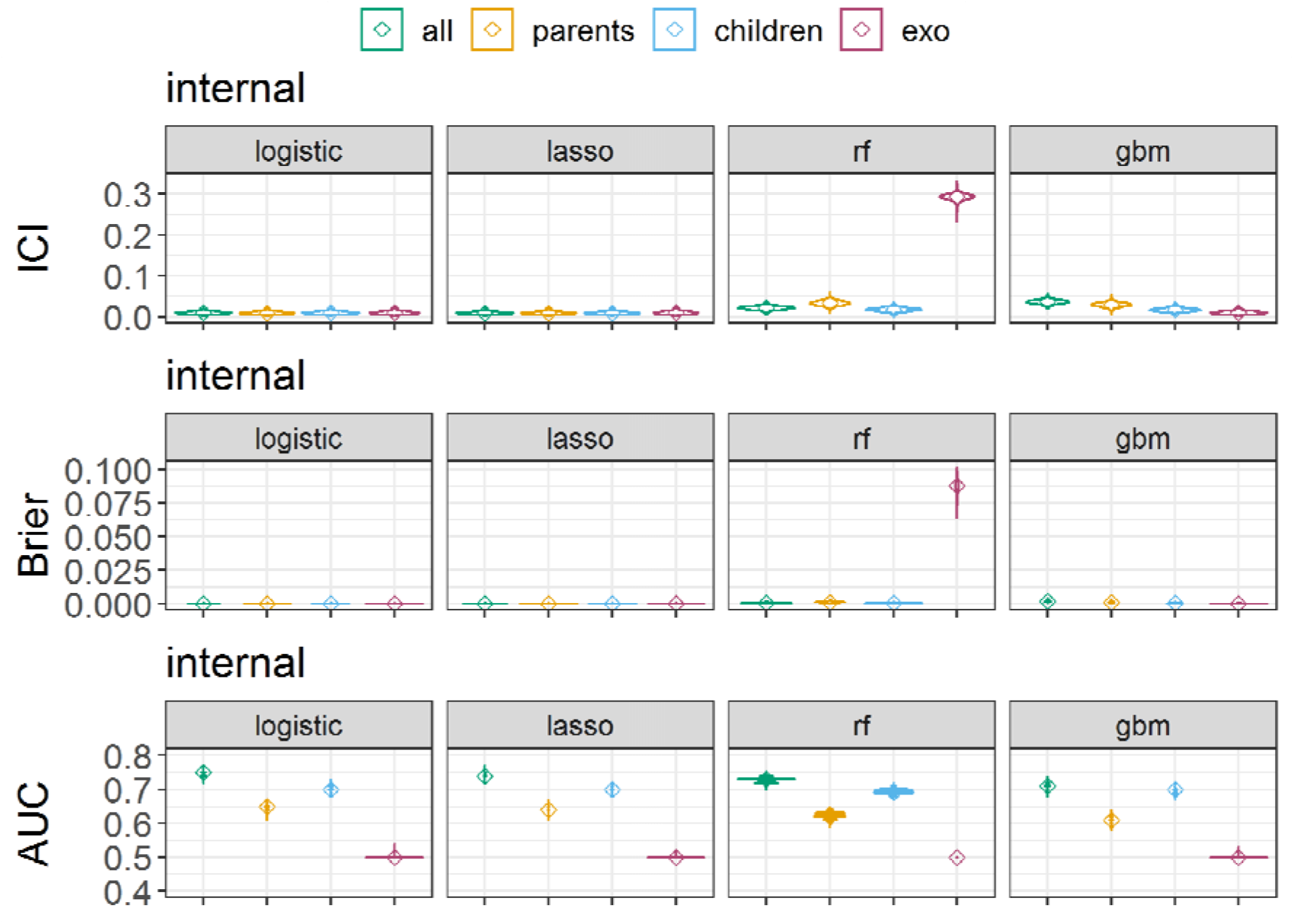
Model performance in the internal validation setting, measured by the integrated calibration index (ICI), Brier score, and area under the receiver operating curve (AUC). Cognitive impairment was predicted using logistic regression, lasso regression, random forest (rf), and generalized boosted regression (gbm) prediction models. Models were trained either with all predictor variables, only parent nodes (direct causes) of the outcome, only children nodes (consequences) of the outcome, or with the exogenous variables age, sex, and APOE ε4 (apoe4). Depicted are the full distributions of ICI, Brier scores, and AUC, smoothed with a Gaussian kernel density function and medians marked with ◊. The displayed metrics were obtained from 10,000 repetitions of data generation and model training on the first imputed dataset.

### Measuring transportability by calibration differences

We compared the transportability of prediction models measured by calibration differences between internal validation and intervention settings (Supplementary Table S6). For this, we focus on logistic regression and lasso regression predicting with all variables, parents, children, and exogenous variables, since they showed good calibration in the internal setting. In all intervention settings, models predicting with parent nodes were more transportable than those predicting with children nodes (Figure 4, Supplementary Figure S3, and Supplementary Figure S4). Models predicting with parents had good transportability in intervention settings, indicated by a similar calibration (thereby small calibration difference) between the internal validation and intervention setting. For example, the median ICI difference between the internal validation and age intervention for logistic regression was very small (ICI -0.009 [-0.045, 0.006]). Models predicting with children variables had low transportability in intervention settings, as indicated by negative calibration differences. For example, logistic regression predicting with children had a median ICI difference between the internal and age intervention setting of -0.300 [-0.322, -0.276], which was 33.3-fold lower than predicting with parents. The largest difference in the median ICI differences between parents and children was in the age intervention setting (logistic regression: 0.291), and the smallest one was in the APOE ε4 setting (logistic regression: 0.008). Logistic regression models predicting with all variables equally or less transportable compared to models predicting with parent variables (e.g., age intervention: all predictors median ICI -0.031 [- 0.073, 0.002], parent predictors: -0.009 [-0.045, 0.006]; tau: all predictors -0.014 [-0.022, - 0.005], parents 0.000 [-0.007, 0.008]). Similar results were observed for the lasso regression models. In the age2 and APOE ε4 intervention setting, the calibration differences were close to zero for logistic and lasso models with all predictors and parent predictors. Using exogenous variables as predictors had calibration differences close to zero, except in the tau intervention setting. We found low transportability of logistic and lasso regression models with all sets of predictors in the tau-intervention setting. Only the models predicting with parent variable had zero calibration differences.

**Figure 4:**
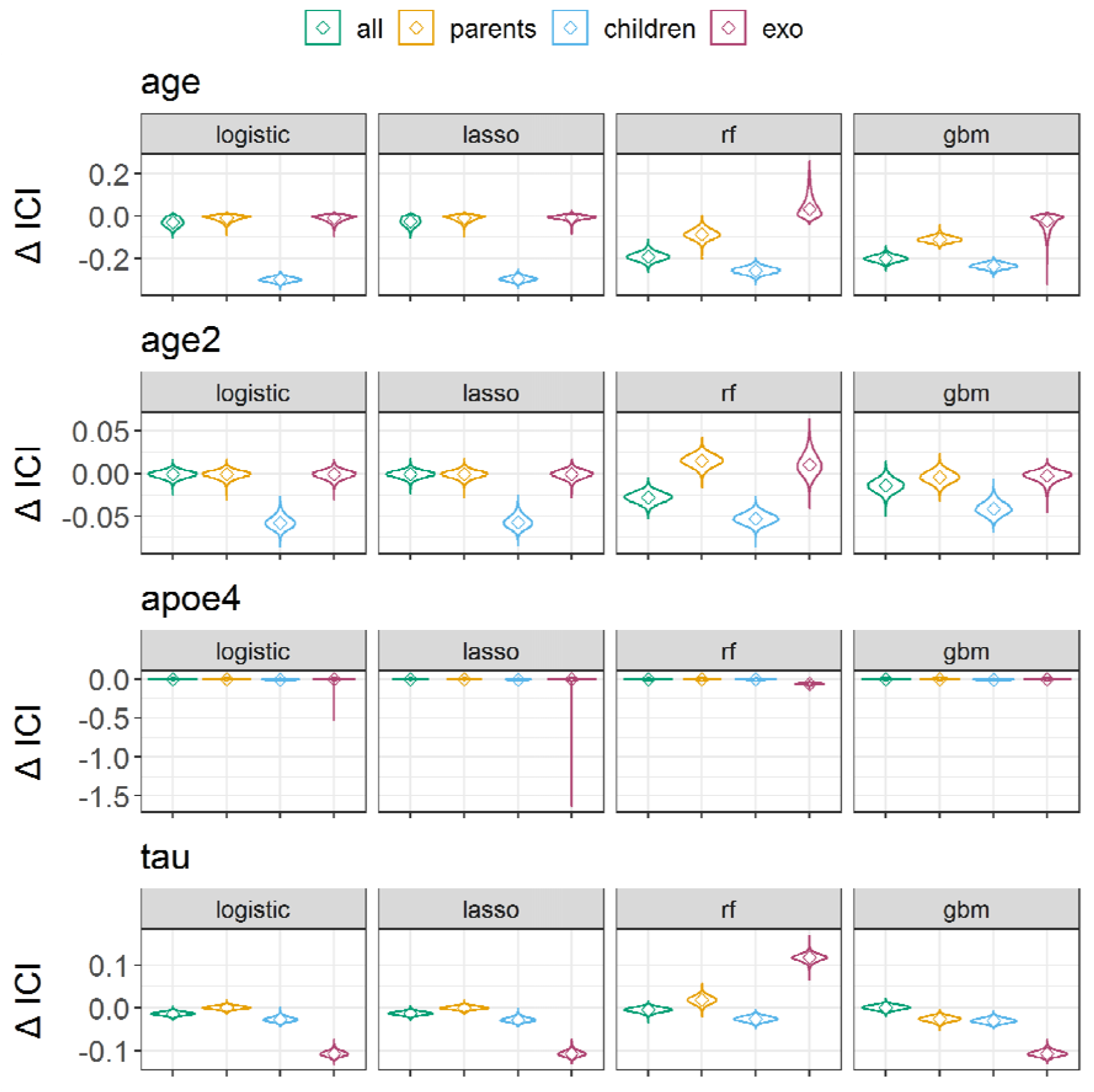
Transportability between internal validation and external settings, measured by the difference of integrated calibration index (ICI). Four intervention test sets were created with 1) reducing the population mean age from 73 to 35 years, 2) reducing the population mean age from 73 to 65 years (age2), 3) reducing the APOE ε4 allele frequency from 46.6% to 5.0%, and 3) changing the SEM-parameters for generating the endogenous variable tau. Cognitive impairment was predicted using logistic regression, lasso regression random forest (rf), and generalized boosted regression (gbm) prediction models. Models were trained either with all predictor variables, only parent nodes (direct causes) of the outcome, only children nodes (consequences) of the outcome, or exogenous variables (exo) age, sex, and APOE ε4 allele frequency. Depicted are the full distributions of ICI differences from 10,000 repetitions on the first imputed dataset, smoothed with a Gaussian kernel density function and medians marked with ◊.

Random forest and GBM models also showed generally lower ICI when using children as predictors compared to parents and higher transportability for parent predictors compared to all predictors. However, random forest and GBM models predicting with parents had ICI differences between internal and external validation differences far from zero. Brier scores supported the same trends as the ICI (Supplementary Figures S4 and S5, Supplementary Table S6).

### Measuring transportability by AUC differences

When measuring transportability with AUC differences, we found inconsistent transportability trends between the intervention settings. In the age-intervention setting, models predicting with children variables also had lower transportability compared to models predicting with parent predictors, indicated by a positive AUC difference between internal validation and intervention setting (logistic regression with parent predictors: -0.01 [-0.04, 0.02], logistic regression with children 0.02 [0.00, 0.04], Figure 5, Supplementary Figures S6 and S7, Supplementary Table S6). In the age2 and tau intervention settings, however, logistic regression models predicting with children increased their AUC by 0.03 compared to the internal validation setting, whereas logistic regression models predicting with parent predictors had AUC differences close to zero. AUC differences indicated similar transportability close to zero for all predictors compared to parent predictors in all intervention settings. Lasso regression, random forest, and GBM models indicated similar trends as logistic regression models.

**Figure 5:**
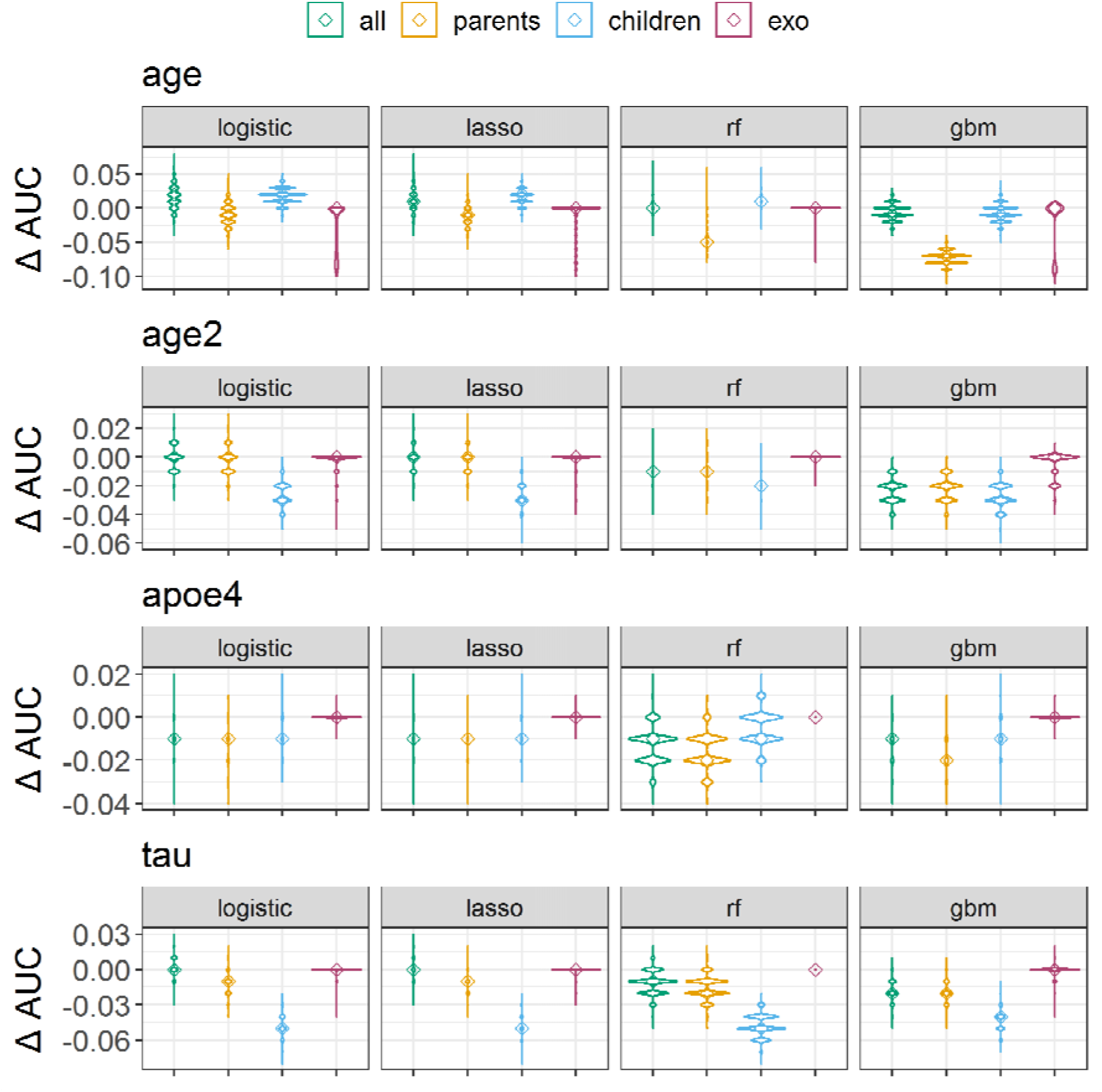
Transportability between internal validation and external settings, measured by the difference of area under the receiver operating curve (AUC). Four intervention test sets were created with 1) reducing the population mean age from 73 to 35 years, 2) reducing the population mean age from 73 to 65 years (age2), 3) reducing the APOE ε4 allele frequency from 46.6% to 5.0%, and 3) changing the SEM-parameters for generating the endogenous variable tau. Cognitive impairment was predicted using logistic regression, lasso regression random forest (rf), and generalized boosted regression (gbm) prediction models. Models were trained either with all predictor variables, only parent nodes (direct causes) of the outcome, only children nodes (consequences) of the outcome, or exogenous variables (exo) age, sex, and APOE ε4 allele frequency. Depicted are the full distributions of AUC differences from 10,000 repetitions on the first imputed dataset, smoothed with a Gaussian kernel density function and medians marked with ◊.

### Sensitivity analyses with other imputed datasets and hyperparameter tuning

We performed sensitivity analysis by using the other two imputed dataset as input for the SEM-model. We did not find any differences between model performance results (ICI, Brier score, and AUC) between replicates that used three different imputed datasets as input (Supplementary Figures S5, S7, S8-9).

Optimizing hyperparameter for random forest models did not meaningfully improve calibration or AUC performance (Supplementary Figure S10). The observed trends for transportability measured by ICI differences (i.e., parent predictors being more transportable compared to children) remained for random forest in the age-interventions settings, but in the tau-intervention setting, optimized random forest models predicting with children may have equal or better ICI transportability (−0.019 [-0.044, 0.012]), compared to parents (−0.032 [-0.063, -0.003]).

## Discussion

In this study, as a first contribution, we have presented a causal data generation approach for assessing the transportability of prediction models for cognitive impairment in synthetic external settings with different distributions of age, APOE ε4 allele frequency, and tau. As a second contribution, we assessed transportability by comparing performance between internal and external validation settings, measured by the discrimination performance (AUC) as in most prior studies,^30–32,34^ but also by calibration (ICI and Brier calibration component).

Both calibration metrics, ICI, and the Brier score confirmed the previous causal theory that prediction models that use direct causes of the outcome for the prediction are generally more transportable.^20,25^ We showed that, under a specific set of interventions, calibration performance remained stable when ML models predicted only with direct causes (parent nodes) but was reduced when predicting with consequences (children nodes) of the outcome ‘cognitive impairment.’ We found that this held true in all prediction models (logistic regression, lasso regression, random forest, and GBM). Measuring transportability by AUC differences indicated inconsistent trends for transportability and did not reflect causal theory. Calibration measures the closeness between the average predicted probabilities and the relative frequency of the outcome event and therefore reflects the actual capability of predicting the state of the outcome variable. In contrast, the AUC depends on the ranking of the predicted risks and measures the probability that a predicted risk for a randomly drawn individual with the event is higher compared to a randomly drawn individual without the event.^40^ With our results, we reinforce the claim that besides discrimination, good calibration is important to achieve clinically useful prediction models.^40^ Using calibration metric differences to measure the transportability of prediction models, however, requires good calibration in the internal setting. It is possible that a model predicting with children variables has better transportability than a model predicting with parent variables if the model with children predictors outperforms the model with parent variables in the internal validation setting.

In our simulation, we found that the random forest and the GBM algorithms were not well calibrated in the internal setting. The miscalibration might have happened because we generated data assuming linear relationships in the SEM, whereas random forest and GBM are designed to capture non-linear relationships.^54^

Our study further showed that causal thinking is essential when selecting predictors for clinical prediction models. Previously developed prediction models for dementia and AD have used brain volumetric measures or cognitive assessment scores as predictors because they reduced prediction errors.^5,55,56^ Similarly to another work^57^, we assumed that these predictors (brain volumetric measures and cognitive test results) are consequences of the cognitive outcome status and therefore predict in the anti-causal direction leading to reduced transportability to external settings. Another work suggested that predictors derived from medical images may often predict in the anti-causal direction as they depict the consequences of a disease, which may raise a caveat towards transportability.^15^

Our application to assess transportability has limitations. First, it cannot be empirically verified if DAGs map causal relationships correctly and if all relevant factors were included. We only included observed variables (other than latent variables for factors), and it is likely that there are unobserved variables involved in the causal process of cognitive impairment. Strong domain expertise is crucial to build accurate DAGs.^43^ Conditional independence tests can test if there is evidence against a given DAG in a dataset.^12^ We applied conditional independence tests to add directed connections between variables, but unexplainable violations were present. One study suggested that causal relationships should generally be assumed to exist between any two variables and that they should only be omitted when evidence is available.^22^ We ensured that our assumptions in the DAG correctly represent the data by using semi-synthetic data so that any possible misspecification of the DAG did not affect the evaluation of the model transportability.

Second, we applied a SEM to the ADNI data to quantify the causal relationships in our DAG. While SEMs are widely applied for this purpose,^16^ their methodology has limitations when using categorical variables.^58–60^ In our application, we had seven categorical variables and found a small correlation between sex and age and between age and APOE ε4. We believe this correlation might stem from biased selection in the ADNI study, which we did not consider in our DAG. Additionally, we found that some SEM parameter estimates were controversial to domain knowledge. For example, the relationship between age and cognitive impairment was estimated to be -0.13, whereas the prevalence of cognitive impairment is known to increase with age. The incorrect SEM estimates may have altered the effects of the interventions. For example, reducing the mean age from 73.9 to 35 only reduced the prevalence of cognitive impairment by 2%. This was likely because the ADNI study included only elderly (>55 years) participants, and the SEM may not have been able to estimate the correct relationship across this limited age range. Third, it was not possible to evaluate our results with real data from external settings. Obtaining observational data from specific healthcare settings for external validation is often difficult due to data protection. Simulating external data may therefore be inevitable for anticipating transportability.^61^ We simulated external validation data by intervening on specific variables (either age or APOE ε4, and tau) at a time. These interventions simplify general distribution shifts between populations in real-world applications where multiple variables can vary jointly. Fourth, our prediction models had suboptimal discrimination performance, similar to other studies,^37^ with the highest achieved AUC of 0.75 in the internal validation setting. Optimizing hyperparameter did not improve the performance. Better performance could be achieved by using multi-modal data and deep neural networks^3,62^.

Our approach to assess the transportability of models predicting cognitive impairment can be extended to overcome the described limitations. Future work could integrate our causal data generation approach with the work of Pölsterl et al. and include unobserved variables in the DAG.^57^ To ensure that the estimated parameters of causal relationships follow biological laws, future refinements could include the SEM with prior distributions as implemented in the *blavaan* R package^63^ or explore alternative latent variable models. Future research could adapt our approach to measure transportability to models using deep learning to other use-cases and high-dimensional data and compare simulated data with real world data (i.e., with a prediction model). For example, recent deep learning models predicted AD from structural brain MR images.^7,64^ Deep learning models, however, often suffer from poor calibration,^65^ whereas we suggested good calibration in the internal validation setting as a requirement for assessing transportability using calibration metrics. New approaches to train calibrated deep learning models offer solutions for better calibration.^66,67^ Additional research to generate synthetic medical images on the basis of causal models has emerged^68^, leaving an exciting open challenge to identify causal structures in complex data^68^ and assess transportability to new settings using interventions and synthetic images.

## Conclusions

Actionable machine learning for health algorithms requires good transportability to new settings but measuring transportability before deployment is challenging. We have used an approach to assess the transportability of prediction models for predicting cognitive impairment using a causal graph and semi-synthetic data to simulate different external validation scenarios. We conclude that calibration metrics may be more suitable than AUC to measure transportability to external settings but require good calibration performance in the internal validation setting. Our results also contain an empirical illustration of the existing theory that models predicting with causes of the outcome have better transportability than those predicting with consequences of the outcome and can help to better select predictors for prediction models. Future research can adapt our approach to other use cases and high-dimensional data such as images and apply new interventions to simulate more realistic external scenarios.

## Data Availability

All data produced are available online at

http://adni.loni.usc.edu

https://tadpole.grand-challenge.org/Data/

## Declarations

This is not a human study. All data used in the study are from public databases. No new human data is generated in the study.

### Ethics, consent and permissions

Ethical review and approval and obtaining informed consent was deemed unnecessary for the present study, because we performed secondary analyses of data obtained from the ADNI study, which was approved by the Institutional Review Boards of all of the participating institutions. All participants represented in the ADNI dataset provided informed written consent as described in. ^41,42^ All methods were carried out in accordance with relevant guidelines and national regulations according to the Declaration of Helsinki (consent for research). More details can be found at adni.loni.usc.edu.

### Consent for publication

Not applicable.

### Availability of data and code

The TADPOLE and ADNI datasets analyzed during the current study are both available at the Alzheimer’s Disease Neuroimaging Initiative (ADNI) database (https://ida.loni.usc.edu) upon consenting to the data sharing agreement. The R code implementing the SEM, data simulation, prediction models and transportability measurements is accessible at https://github.com/JanaFe/AssessingTransportability.

### Author’s Contributions

MP and SK conceptualized the study. JF implemented the methods in R, ran the analyses, and drafted a first version of the manuscript. JF, MP, SK interpreted the results. SK supervised the study. TK consulted the creation of the DAG. All authors provided critical input to the manuscript and approved the final version.

## Acknowledgements

Data collection and sharing for this project was funded by the Alzheimer’s Disease Neuroimaging Initiative (ADNI) (National Institutes of Health Grant U01 AG024904) and DOD ADNI (Department of Defense award number W81XWH-12-2-0012). ADNI is funded by the National Institute on Aging, the National Institute of Biomedical Imaging and Bioengineering, and through generous contributions from the following: AbbVie, Alzheimer’s Association; Alzheimer’s Drug Discovery Foundation; Araclon Biotech; BioClinica, Inc.; Biogen; Bristol-Myers Squibb Company; CereSpir, Inc.; Cogstate; Eisai Inc.; Elan Pharmaceuticals, Inc.; Eli Lilly and Company; EuroImmun; F. Hoffmann-La Roche Ltd and its affiliated company Genentech, Inc.; Fujirebio; GE Healthcare; IXICO Ltd.; Janssen Alzheimer Immunotherapy Research & Development, LLC.; Johnson & Johnson Pharmaceutical Research & Development LLC.; Lumosity; Lundbeck; Merck & Co., Inc.; Meso Scale Diagnostics, LLC.; NeuroRx Research; Neurotrack Technologies; Novartis Pharmaceuticals Corporation; Pfizer Inc.; Piramal Imaging; Servier; Takeda Pharmaceutical Company; and Transition Therapeutics. The Canadian Institutes of Health Research is providing funds to support ADNI clinical sites in Canada. Private sector contributions are facilitated by the Foundation for the National Institutes of Health (www.fnih.org). The grantee organization is the Northern California Institute for Research and Education, and the study is coordinated by the Alzheimer’s Therapeutic Research Institute at the University of Southern California. ADNI data are disseminated by the Laboratory for Neuro Imaging at the University of Southern California.

## Footnotes

1 https://worldpopulationreview.com/country-rankings/median-age

