## Supplementary Material for "Assessing the transportability of clinical prediction models for cognitive impairment using causal models"

### Supplementary Text 1: Details on data processing

We obtained age, sex, education and the presence of the APOE ε4 genotype (0 or 1). TADPOLE includes brain volumetric measurements which were extracted from structural MRI images using an atlas-based technique. We included hippocampal, fusiform, midtemporal lobe, ventricle and intracranial volume (ICV) measurements. Reduced $^{18}$F-fluorodeoxyglucose-positron emission tomography (FDG-PET) highlights brain cells with reduced metabolism as a sign of neurodegeneration [1] and the FDG-PET measurement was obtained from TADPOLE. Further selected biomarkers include measured tau and amyloid-beta proteins in the cerebrospinal fluid (CSF-Tau, CSF-Aβ). The history of smoking and alcoholism for each individual was obtained from ADNI MEDHIST and merged with the TADPOLE data. Body weight and height measurements for the individuals at baseline were obtained from ADNI VITALS to calculate body mass index (BMI). We defined a history of hypertension (yes or no) if an individual had a systolic blood pressure greater than 140 mmHg or diastolic greater than 90 mmHg at baseline in ADNI VITALs or if a history of hypertension was described in ADNI HMHYPERT. History of cardiovascular disease (yes or no) was obtained from ADNI MEDHIST. We obtained Mini Mental state exam (mmse) scores that rate cognitive abilities from 0-30. Diagnoses in the ADNI dataset were recorded as mutually exclusive categories: either cognitive normal (n=417, 24.0%), subjective memory complaint (n=106, 6.1%), early (n=310, 17.9%) or late mild cognitive impairment (n=562, 32.4%), and Alzheimer's Disease (n=342, 19.7%). We chose cognitive impairment as outcome variable, defined as having mild cognitive impairment or Alzheimer's disease.

### Supplementary Text 2: Details on added causal edges

We tested if the DAG fulfils the criteria of conditional independence with respect to the obtained ADNI dataset. P values from conditional independence testing were adjusted according to the number of tests. We reviewed connections that had violated conditional independence (P value adjusted with Bonferroni correction for multiple testing <0.05) and added the following connections.

- Apoe → hippocampus
- Age → icv
- Sex → icv
- Sex → fdg
- Sex → cardio
- Sex → cognitive status
- BMI → Hippocampus
- BMI → cognitive status
- Educ → cardio
- Educ → hypertension
- Tau → hippocampus
- Tau → mmse
- Aβ → mmse

Note: Apoe = prevalence of APOE ε4 gene, icv = intracranial volume, fdg = Fluorodeoxyglucose positron emission tomography, cardio = history of cardiovascular events, Educ = education, Tau = tau-protein measured in cerebrospinal fluid, Aβ = Aβ-protein measured in cerebrospinal fluid, mmse = minimental state exam.

### Supplementary Tables

Table S1: Scientific evidence of assumed causal relationships. These references document scientific evidence that was used to map causal relationships depicted as arrows in the directed acyclic graph. Listed are arrows, quotes from scientific publications and the respective reference. *Aβ: Amyloid β; †AD: Alzheimer's Disease, ‡mmse: Mini Mental State Exam score

| Arrow | Statement | Source |
|---|---|---|
| APOE ε4 → Aβ | "The effect of APOE ε4 in driving amyloid pathology is likely to outweigh the protective effect of APOE2." | Yamazaki Y, Zhao N, Caulfield TR, Liu CC, Bu G. Apolipoprotein E and Alzheimer disease: pathobiology and targeting strategies. *Nat Rev Neurol*. 2019;15(9):501-518. doi:10.1038/s41582-019-0228-7 |
| APOE ε4 → Tau | "ApoE affects tau pathogenesis, neuroinflammation, and tau-mediated neurodegeneration independently of amyloid-β pathology" | Shi Y, Yamada K, Liddelow SA, Smith ST, Zhao L, Luo W, Tsai RM, Spina S, Grinberg LT, Rojas JC, Gallardo G, Wang K, Roh J, Robinson G, Finn MB, Jiang H, Sullivan PM, Baufeld C, Wood MW, Sutphen C, McCue L, Xiong C, Del-Aguila JL, Morris JC, Cruchaga C; Alzheimer's Disease Neuroimaging Initiative, Fagan AM, Miller BL, Boxer AL, Seeley WW, Butovsky O, Barres BA, Paul SM, Holtzman DM. APOE ε4 markedly exacerbates tau-mediated neurodegeneration in a mouse model of tauopathy. Nature. 2017 Sep 28;549(7673):523-527. doi: 10.1038/nature24016. Epub 2017 Sep 20. PMID: 28959956; PMCID: PMC5641217. |
| APOE ε4 → hippocampus volume | "Hippocampus may be particularly vulnerable to further degeneration in APOE ε4 carriers as they enter middle and old age" | O'Dwyer L, Lamberton F, Matura S, et al. Reduced hippocampal volume in healthy young APOE ε4 carriers: an MRI study. *PLoS One*. 2012;7(11):e48895. doi:10.1371/journal.pone.0048895 |
| Aβ deposits → medial temporal atrophy, synaptic dysfunction and cognitive decline. | "AD† begins with Aβ accumulation in the brain, which ultimately leads to synaptic dysfunction, neurodegeneration, and cognitive or functional decline. Neurodegeneration is detected | Weiner MW, Aisen PS, Jack CR Jr, et al. The Alzheimer's disease neuroimaging initiative: progress report and future plans. *Alzheimers Dement*. 2010;6(3):202-11.e7. doi:10.1016/j.jalz.2010.03.007 |

| | | |
|---|---|---|
| | by a rise of CSF tau species, synaptic dysfunction (measured by FDG-PET), and neuron loss indicated by atrophy, most notably in medial temporal lobe (measured with MRI)” | |
| cardio → development/progression of AD† (using CSF-Aβ as a proxy) | “Several studies have identified modifiable cardiometabolic diseases including type 2 diabetes, metabolic syndrome, obesity, and other cardiovascular risk factors for the development and progression of AD†.” | Barnes DE, Yaffe K. The projected effect of risk factor reduction on Alzheimer’s disease prevalence. Lancet Neurol. 2011;10:819–28. [PMC free article] |
| Sex → AD† (proxy: cognitive status) | “More than 5.5 million Americans, including an estimated 5.3 million people 65 years and older, are currently living with AD† dementia; approximately two-thirds of whom are women.<br>Debate: New research is needed to understand whether the relationship between cardiometabolic factors and risk of AD† differs by sex, and how this difference varies by age” | Nebel RA, Aggarwal NT, Barnes LL, et al. Understanding the impact of sex and gender in Alzheimer's disease: A call to action. *Alzheimers Dement*. 2018;14(9):1171-1183. doi:10.1016/j.jalz.2018.04.008 |
| Sex → hippocampal volume | Found absolute hippocampal volumes were larger in men than women, but suggest that gender differences may be due to headsize or pre-processing. | Gabor Perlaki, Gergely Orsi, Eniko Plozer, Anna Altbacker, Gergely Darnai, Szilvia Anett Nagy, Reka Horvath, Arnold Toth, Tamas Doczi, Norbert Kovacs, Peter Bogner, Attila Schwarcz, Jozsef Janszky. Are there any gender differences in the hippocampus volume after head-size correction? A volumetric and voxel-based morphometric study. Neuroscience Letters 570, 2014,<br>Pages 119-123, https://doi.org/10.1016/j.neulet.2014.04.013. |
| Sex → icv | “few sex differences (if any) remain statistically significant, and their size is quite reduced” | Sanchis-Segura C, Ibañez-Gual MV, Adrián-Ventura J, Aguirre N, Gómez-Cruz ÁJ, Avila C, Forn C. Sex differences in gray matter volume: how many and how large are they really? Biol Sex Differ. 2019 Jul 1;10(1):32. doi: 10.1186/s13293-019-0245-7. |

| | | |
|---|---|---|
| | | PMID: 31262342; PMCID: PMC6604149. |
| Sex → FDG brain metabolism (proxy: FDG-PET) | “Report discloses different sex- and age-related brain metabolism.” | Hsieh TC, Lin WY, Ding HJ, Sun SS, Wu YC, Yen KY, Kao CH. Sex- and age-related differences in brain FDG metabolism of healthy adults: an SPM analysis. J Neuroimaging. 2012 Jan;22(1):21-7. doi: 10.1111/j.1552-6569.2010.00543.x. Epub 2011 Feb 18. PMID: 21332873. |
| Age → intracranial volume (icv) | “Human intracranial volume does not stay constant during adulthood but instead shows a small increase during young adulthood and a decrease thereafter from the fourth decade of life.” | Caspi Y, Brouwer RM, Schnack HG, van de Nieuwenhuijzen ME, Cahn W, Kahn RS, Niessen WJ, van der Lugt A, Hulshoff Pol H. Changes in the intracranial volume from early adulthood to the sixth decade of life: A longitudinal study. biorxiv, 2020. https://doi.org/10.1101/677898 |
| Age → brain atrophy | “With old age there is an overall reduction in brain volume with the majority of grey matter shrinkage occurring in the hippocampus and prefrontal cortex (PFC)“ | Hou, Y., Dan, X., Babbar, M. *et al.* Ageing as a risk factor for neurodegenerative disease. *Nat Rev Neurol* **15,** 565–581 (2019). https://doi.org/10.1038/s41582-019-0244-7 |
| Bmi → hippocampus volume | “Being overweight is associated with hippocampal atrophy.” | Cherbuin, N., Sargent-Cox, K., Fraser, M. *et al.* Being overweight is associated with hippocampal atrophy: the PATH Through Life Study. *Int J Obes* **39,** 1509–1514 (2015). https://doi.org/10.1038/ijo.2015.106 |
| Aβ deposits (proxy CSF-Aβ) → brain atrophy (hippocampus, ventricles) | “The mechanisms that link amyloid-β to neurodegeneration are poorly understood and perplexing. CSF measures of amyloid-$\beta_{1-42}$ have also been associated with whole brain atrophy. However, not all studies have found such relationships in cross-sectional data. In the Alzheimer’s Disease Neuroimaging Initiative (ADNI), no relationship was detected | Jagust W. Is amyloid-β harmful to the brain? Insights from human imaging studies. *Brain*. 2016;139(Pt 1):23-30. doi:10.1093/brain/awv326 |

| | | |
|---|---|---|
| | between brain amyloid-β and regional brain atrophy in 280 normal older people, 30% of whom had evidence of brain amyloid deposition although evidence for such associations were seen in those with MCI.” | |
| Tau-deposits (proxy: CSF-Tau) → brain atrophy | “While elevated CSF tau concentrations have been shown to be associated with lower grey matter density (GMD) in AD†-specific regions, this correlation has yet to be examined for plasma in a large study. Plasma tau may serve as a non-specific marker for neurodegeneration but is still relevant to AD† considering low GMD was associated with plasma tau in Aβ+ participants and not Aβ-participants.” | Deters KD, Risacher SL, Kim S, et al. Plasma Tau Association with Brain Atrophy in Mild Cognitive Impairment and Alzheimer's Disease. *J Alzheimers Dis*. 2017;58(4):1245-1254. doi:10.3233/JAD-161114 |
| Bmi → brain atrophy (proxy: icv, hippocampus, ventricles) | “Adiposity in early-old age is related to reduced global gray matter later in life in this diverse sample. Future studies are warranted to elucidate causal relationships and explore region-specific associations.” | Caunca MR, Gardener H, Simonetto M, Cheung YK, Alperin N, Yoshita M, DeCarli C, Elkind MSV, Sacco RL, Wright CB, Rundek T. Measures of obesity are associated with MRI markers of brain aging: The Northern Manhattan Study. Neurology. 2019 Aug 20;93(8):e791-e803. doi: 10.1212/WNL.0000000000007966. Epub 2019 Jul 24. PMID: 31341005; |
| | “Overall, our results suggest that midlife obesity may be an important modifier of brain atrophy in individuals who are developing cognitive impairment and dementia, while it has little effect on structural brain integrity in nondemented older adults.” | Driscoll I, Beydoun MA, An Y, et al. Midlife obesity and trajectories of brain volume changes in older adults. *Hum Brain Mapp*. 2012;33(9):2204-2210. doi:10.1002/hbm.21353 |
| Bmi → cognitive impairment | “Lower baseline BMI was associated with more rapid cognitive decline in MCI.” | Cronk BB, Johnson DK, Burns JM; Alzheimer's Disease Neuroimaging Initiative. Body mass index and cognitive decline in mild cognitive impairment. Alzheimer Dis Assoc Disord. 2010 Apr-Jun;24(2):126-30. doi: 10.1097/WAD.0b013e3181a6bf3f. PMID: 19571736; PMCID: |

| | | |
|---|---|---|
| | "BMI-defined overweight, but not obesity, was associated with a lower risk of cognitive impairment among elderly Chinese adults, while large weight loss was associated with an increased risk." | PMC3068614.<br><br>Wu S, Lv X, Shen J, Chen H, Ma Y, Jin X, Yang J, Cao Y, Zong G, Wang H, Yuan C. Association between body mass index, its change and cognitive impairment among Chinese older adults: a community-based, 9-year prospective cohort study. Eur J Epidemiol. 2021 Aug 9. doi: 10.1007/s10654-021-00792-y. Epub ahead of print. PMID: 34370136. |
| Alcohol abuse → ventricular volume | "Compared to control subjects, alcohol patients showed bilaterally decreased prefrontal lobes (11% reduction) and increased lateral ventricles (up to 42% enlargement)." | Wobrock T, Falkai P, Schneider-Axmann T, Frommann N, Wölwer W, Gaebel W. Effects of abstinence on brain morphology in alcoholism: a MRI study. *Eur Arch Psychiatry Clin Neurosci*. 2009;259(3):143-150. doi:10.1007/s00406-008-0846-3 |
| Tau-deposits (proxy: CSF-Tau) → hippocampus volume | "The CSF tau and P-tau levels, but not the CSF $A\beta_{42}$ level, correlated with HV, suggesting that CSF tau markers reflect the neuronal loss associated with the physiopathological process of AD[†]." | Leonardo C. de Souza, Marie Chupin, Foudil Lamari, Claude Jardel, Delphine Leclercq, Olivier Colliot, Stéphane Lehéricy, Bruno Dubois, Marie Sarazin,<br>CSF tau markers are correlated with hippocampal volume in Alzheimer's disease,<br>Neurobiology of Aging,<br>Volume 33, Issue 7,<br>2012,<br>Pages 1253-1257,<br>ISSN 0197-4580,<br>https://doi.org/10.1016/j.neurobiolaging.2011.02.022. |
| Alcohol abuse → hippocampus volume | "Both alcoholic men and women had significantly smaller right hippocampi and larger cerebrospinal fluid volumes than healthy subjects of the same sex." | Agartz I, Momenan R, Rawlings RR, Kerich MJ, Hommer DW. Hippocampal volume in patients with alcohol dependence. Arch Gen Psychiatry. 1999 Apr;56(4):356-63. doi: 10.1001/archpsyc.56.4.356. PMID: 10197833.0197833/ |
| age → synaptic dysfunction (proxy: fdg-PET) | Aging brains exhibit neuroinflammation | Barrientos RM, Kitt MM, Watkins LR, Maier SF. Neuroinflammation in the normal aging hippocampus. |

| | | |
|---|---|---|
| | | Neuroscience. 2015 Nov 19;309:84-99. doi: 10.1016/j.neuroscience.2015.03.007. Epub 2015 Mar 12. PMID: 25772789; PMCID: PMC4567963. |
| CSF-Aβ → synaptic dysfunction (proxy: FDG-PET) | "Aβ oligomers were potent CNS neurotoxins that rapidly inhibited long-term potentiation and, with time, caused selective nerve cell death. The mechanism was attributed to disrupted signaling involving the tyrosine-protein kinase Fyn, mediated by an unknown toxin receptor." | Lambert MP, Barlow AK, Chromy BA, et al. Diffusible, nonfibrillar ligands derived from Aβ1-42 are potent central nervous system neurotoxins. *Proc Natl Acad Sci U S A*. 1998;95(11):6448-6453. doi:10.1073/pnas.95.11.6448 |
| education → cognitive reserve (proxy: FDG-PET) | "Symptoms of AD[†] are recognized later among those with less education."<br><br>"The CR hypothesis postulates that CR reduces the prevalence and incidence of AD[†]. It also hypothesizes that among those who have greater initial cognitive reserve (in contrast to those with less reserve) greater brain pathology occurs before the clinical symptoms of disease manifest." | Roe CM, Xiong C, Grant E, Miller JP, Morris JC. Gen *Arch Neurol*. 2008;65(1):108-111. doi:10.1001/archneurol.2007.11<br><br>Meng X, D'Arcy C. Education and dementia in the context of the cognitive reserve hypothesis: a systematic review with meta-analyses and qualitative analyses. *PLoS One*. 2012;7(6):e38268. doi:10.1371/journal.pone.0038268 |
| education → cognitive status (proxy: Mini Mental State Exam, MMSE) | "The risk of Alzheimer's disease associated with subjective memory complaints was higher in highly educated persons than in persons with a low education. In highly educated persons without objective cognitive impairment (Mini-Mental State Examination score, 29 or 30) the risk of Alzheimer's disease was highest (age- and sex-adjusted hazard ratio, 2.98; 95% CI,1.76 –5.02)."<br><br>"Lower education was associated with a greater risk for dementia in many but not all studies. A relationship between education and risk for dementia | van Oijen, M., de Jong, F.J., Hofman, A., Koudstaal, P.J. and Breteler, M.M.B. (2007), Subjective memory complaints, education, and risk of Alzheimer's disease. Alzheimer's & Dementia, 3: 92-97. https://doi.org/10.1016/j.jalz.2007.01.011https://doi.org/10.1016/j.jalz.2007.01.011<br><br>Sharp ES, Gatz M. Relationship between education and dementia: an updated systematic review. *Alzheimer Dis Assoc Disord*. 2011;25(4):289-304. |

| | | |
|---|---|---|
| | was more consistent in developed compared to developing regions. Age, gender, race/ethnicity, and geographical region moderated the relationship” | doi:10.1097/WAD.0b013e318211c83c |
| education → cognitive test performance (proxy: MMSE) | “educational level has been highly related to performance on neuropsychological testing. Education quality, which can be captured through literacy measures, has been found to have a greater influence on certain neuropsychological test performance than age and may also delay the onset of identifiable cognitive deficits among those with brain diseases”<br><br>“People with more education have lower prevalence of dementia, more years of cognitively healthy life, and fewer years with dementia.” | Ardila, A. (2007). The impact of culture on neuropsychological test performance. *International handbook of cross-cultural neuropsychology in International handbook of cross-cultural neuropsychology* (pp. 23–44). Lawrence Erlbaum Associates, Inc.<br><br>Crimmins EM, Saito Y, Kim JK, Zhang YS, Sasson I, Hayward MD. Educational Differences in the Prevalence of Dementia and Life Expectancy with Dementia: Changes from 2000 to 2010. J Gerontol B Psychol Sci Soc Sci. 2018 Apr 16;73(suppl_1):S20-S28. doi: 10.1093/geronb/gbx135. PMID: 29669097; PMCID: PMC6019027. |
| Aβ and tau deposits (proxy: CSF-Aβ and CSF-Tau) → cognitive test performance (proxy: mmse‡) | “CSF Aβ43 was demonstrated to be significantly reduced in patients already by the time that aMCI or AD[†] was diagnosed, compared to controls, and this change must have occurred during the preclinical period. Since our results suggested that CSF Aβ43 distinguishes between subgroups of patients with aMCI better than CSF Aβ42, it may prove to be a useful additional biomarker for identifying aMCI patients at greatest risk of AD[†].” | Lauridsen C, Sando SB, Shabnam A, et al. Cerebrospinal Fluid Levels of Amyloid Beta 1-43 in Patients with Amnestic Mild Cognitive Impairment or Early Alzheimer's Disease: A 2-Year Follow-Up Study. *Front Aging Neurosci*. 2016;8:30. Published 2016 Mar 1. doi:10.3389/fnagi.2016.00030<br><br>Li G, Sokal I, Quinn JF, Leverenz JB, Brodey M, Schellenberg GD, Kaye JA, Raskind MA, Zhang J, Peskind ER, Montine TJ. CSF tau/Aβ42 ratio for increased risk of mild cognitive impairment: a follow-up study. Neurology. 2007 Aug 14;69(7):631-9. doi: 10.1212/01.wnl.0000267428.62582.aa. PMID: 17698783. |

| | | |
|---|---|---|
| Smoking → hypertension | "Effect of smoking on blood pressure" | Seltzer CC. Effect of smoking on blood pressure. Am Heart J. 1974 May;87(5):558-64. doi: 10.1016/0002-8703(74)90492-x. PMID: 4818700. https://pubmed.ncbi.nlm.nih.gov/4818700/ |
| Alcohol abuse → hypertension | "In alcohol-consuming populations, the amount of alcohol consumption has significant impact on blood pressure values, the prevalence of hypertension" | Huntgeburth M, Ten Freyhaus H, Rosenkranz S. Alcohol consumption and hypertension. Curr Hypertens Rep. 2005 Jun;7(3):180-5. doi: 10.1007/s11906-005-0007-2. PMID: 15913491. |
| AD dementia → FDG-PET | "Patients with AD† dementia show a characteristic pattern of hypometabolism in precuneus, posterior cingulate, parietal cortices, lateral temporal cortex, frontal cortices, and medial temporal lobe, that can be measured by Fluorodeoxyglucose (FDG) PET imaging." | Alexander GE, Chen K, Pietrini P, Rapoport SI, Reiman EM. Longitudinal PET evaluation of cerebral metabolic decline in dementia: a potential outcome measure in Alzheimer's disease treatment studies. Am J Psychiatry. 2002;159:738–45. |

Table S2: Estimates of assumed causal relationships obtained by structural equation modelling (SEM). The input dataset for the SEM-model was obtained from the Alzheimer's Disease Neuroimaging Initiative (ADNI) dataset. Missing values in the dataset were imputed three times. Each of the three imputed datasets were used as input to the SEM. Listed are the SEM parameters estimates obtained from each of the three imputed datasets (Estimate 1, Estimate 2, and Estimate 3).

| | Estimate 1 | Estimate 2 | Estimate 3 |
|---|---|---|---|
| **Estimated coefficients** | | | |
| educ~ s | 0.38 | 0.38 | 0.38 |
| educ~ a | -0.10 | -0.10 | -0.10 |
| bmi~ a | -0.15 | -0.15 | -0.15 |
| bmi~ s | 0.15 | 0.15 | 0.14 |
| bmi~ educ | -0.08 | -0.08 | -0.08 |
| alc~ a | -0.45 | -0.45 | -0.44 |
| alc~ s | -0.45 | -0.40 | -0.42 |
| alc~ educ | -0.07 | -0.09 | -0.09 |
| smo~ a | -0.06 | -0.06 | -0.06 |
| smo~ s | 0.05 | 0.05 | 0.08 |
| smo~ educ | 0.02 | 0.00 | -0.01 |
| hyp~ a | 0.11 | 0.10 | 0.10 |
| hyp~ s | 0.09 | 0.11 | 0.12 |
| hyp~ bmi | 0.01 | 0.00 | 0.01 |
| hyp~ alc | -0.17 | -0.19 | -0.18 |
| hyp~ smo | -0.54 | -0.54 | -0.58 |

| | | | |
|---|---|---|---|
| hyp~ educ | -0.13 | -0.15 | -0.16 |
| cardio~ a | 0.13 | 0.13 | 0.13 |
| cardio~ s | 0.14 | 0.13 | 0.13 |
| cardio~ bmi | 0.20 | 0.20 | 0.20 |
| cardio~ educ | -0.03 | -0.03 | -0.03 |
| cardio~ alc | 0.04 | 0.04 | 0.02 |
| cardio~ smo | 0.20 | 0.22 | 0.17 |
| cardio~ hyp | 0.43 | 0.44 | 0.41 |
| ab~ a | -0.14 | -0.15 | -0.14 |
| ab~ ap | -0.84 | -0.84 | -0.80 |
| tau~ a | 0.37 | 0.38 | 0.37 |
| tau~ ap | 0.57 | 0.52 | 0.53 |
| tau~ alc | 0.57 | 0.58 | 0.55 |
| tau~ hyp | 0.14 | 0.14 | 0.12 |
| dx~ a | -0.13 | -0.14 | -0.14 |
| dx~ ap | 0.01 | 0.01 | 0.02 |
| dx~ s | 0.56 | 0.55 | 0.56 |
| dx~ educ | -0.12 | -0.10 | -0.12 |
| dx~ ab | -0.32 | -0.34 | -0.31 |
| dx~ tau | 0.41 | 0.44 | 0.45 |
| dx~ cardio | 0.05 | 0.04 | 0.05 |
| dx~ bmi | -0.10 | -0.10 | -0.10 |
| fdg~ a | 0.11 | 0.17 | 0.12 |
| fdg~ s | 0.08 | 0.00 | 0.03 |
| fdg~ dx | -0.31 | -0.29 | -0.30 |
| fdg~ ab | 0.23 | 0.25 | 0.23 |
| fdg~ tau | -0.36 | -0.40 | -0.38 |
| fdg~ alc | 0.37 | 0.43 | 0.40 |
| fdg~ smo | 0.03 | 0.04 | 0.02 |
| fdg~ educ | 0.06 | 0.05 | 0.04 |
| fdg~ hyp | -0.01 | 0.01 | -0.03 |
| fdg~ cardio | 0.05 | 0.03 | 0.04 |
| hi~ a | -0.19 | -0.15 | -0.17 |
| hi~ ap | -0.04 | 0.01 | 0.00 |
| hi~ s | 0.56 | 0.52 | 0.53 |
| hi~ bmi | 0.09 | 0.09 | 0.10 |
| hi~ dx | -0.27 | -0.23 | -0.25 |
| hi~ ab | 0.15 | 0.17 | 0.17 |
| hi~ alc | 0.34 | 0.38 | 0.36 |
| hi~ tau | -0.37 | -0.46 | -0.41 |
| ve~ dx | 0.34 | 0.34 | 0.33 |
| ve~ ab | -0.14 | -0.13 | -0.16 |
| ve~ alc | -0.52 | -0.54 | -0.54 |
| icv~ a | 0.00 | 0.01 | 0.01 |
| icv~ s | 1.03 | 1.01 | 1.02 |
| icv~ dx | 0.17 | 0.19 | 0.17 |

| | | | |
|---|---|---|---|
| icv~ ab | -0.03 | -0.03 | -0.04 |
| icv~ tau | -0.25 | -0.30 | -0.28 |
| me~ dx | -0.41 | -0.40 | -0.39 |
| me~ educ | 0.11 | 0.12 | 0.12 |
| me~ a | -0.04 | -0.05 | -0.04 |
| me~ ab | 0.02 | 0.05 | 0.04 |
| me~ tau | 0.03 | 0.00 | 0.01 |
| me~ fdg | 0.25 | 0.22 | 0.25 |
| me~ ve | 0.05 | 0.03 | 0.05 |
| me~ hi | 0.18 | 0.17 | 0.17 |
| me~ icv | 0.05 | 0.05 | 0.04 |
| **Thresholds for categorization** | | | |
| alc | 2.22 | 2.21 | 2.23 |
| smo | 0.76 | 0.77 | 0.74 |
| hyp | 0.53 | 0.52 | 0.53 |
| cardio | -0.31 | -0.30 | -0.30 |
| dx | -0.30 | -0.32 | -0.30 |
| **Intercept estimates** | | | |
| educ | -0.14 | -0.14 | -0.14 |
| bmi | 0.04 | 0.04 | 0.05 |
| ab | 0.47 | 0.48 | 0.45 |
| tau | -0.15 | -0.16 | -0.15 |
| fdg | 0.06 | 0.10 | 0.07 |
| hi | -0.17 | -0.19 | -0.18 |
| ve | -0.22 | -0.22 | -0.22 |
| icv | -0.62 | -0.62 | -0.62 |
| me | 0.15 | 0.13 | 0.15 |
| **Variance estimates** | | | |
| educ | 0.96 | 0.96 | 0.96 |
| bmi | 0.96 | 0.96 | 0.96 |
| alc | 1 | 0.99 | 0.99 |
| smo | 1 | 1.00 | 1.00 |
| hyp | 0.98 | 0.98 | 0.98 |
| cardio | 0.95 | 0.95 | 0.95 |
| ab | 0.80 | 0.80 | 0.81 |
| tau | 0.52 | 0.50 | 0.54 |
| dx | 0.80 | 0.78 | 0.78 |
| fdg | 0.58 | 0.56 | 0.57 |
| hi | 0.49 | 0.47 | 0.47 |
| ve | 0.43 | 0.40 | 0.41 |
| icv | 0.61 | 0.59 | 0.60 |
| me | 0.44 | 0.46 | 0.45 |

Table S3: Estimates of parameters obtained from the structural equation model (SEM). For all endogenous variables, SEM estimates intercepts, slope and residual variance for linear relationships between variables, specified in the DAG. SEM was applied three times on three imputed datasets. The table presents the obtained SEM-parameters on the first imputed dataset. For categorical variables, a latent variable and its threshold is estimated to divide the data into categories (assigned as 1, if the value exceeds the threshold). The included variables are: APOE ε4 (apoe4), age, sex, education (education), CSF-Aβ (aβ), history of alcohol abuse (alcohol), history of smoking behavior (smoking), Body Mass Index (bmi), history of hypertension (hypertension), CSF-tau (tau), history of cardiovascular events (cardio), cognitive status (cogn), hippocampus (hippocampus), ventricles (ventricles), intracranial volume (icv), FDG-PET (fdg), Mini-Mental State Exam score (mmse).

| Variable | Linear equation | Variance | Threshold |
|---|---|---|---|
| education | -0.14 + 0.38 * sex -0.10 * age | 0.96 | |
| bmi | 0.04 - 0.15 * age + 0.15 * sex - 0.08 * education | 0.96 | |
| alcohol | -0.45 * age - 0.45 * sex - 0.07 * education | 1.00 | 2.22 |
| smoking | -0.06 * age + 0.05 * sex + 0.02 * education | 1.00 | 0.76 |
| hypertension | 0.11 * age + 0.09 * sex + 0.01 * bmi - 0.17 * alcohol - 0.54 * smoking - 0.13 * education | 0.98 | 0.53 |
| cardio | 0.13 * age + 0.14 * sex + 0.20 * bmi - 0.03 * education + 0.04 * alcohol + 0.20 * smoking + 0.43 * hypertension | 0.95 | -0.31 |
| aβ | 0.47 - 0.14 * age - 0.84 * apoe4 | 0.80 | |
| tau | -0.15 + 0.37 * age + 0.57 * apoe4 + 0.57 * alcohol + 0.14 * hypertension | 0.52 | |
| cognitive impairment (cogn) | -0.13* age + 0.01 * apoe4 + 0.56 * sex - 0.12 * education - 0.32 * aβ + 0.41 * tau + 0.045 * cardio - 0.10 * bmi | 0.80 | -0.30 |
| fdg | 0.06 + 0.11 * age + 0.08 * sex - 0.31 * cogn + 0.23 * aβ - 0.36 * tau + 0.37 * alc + 0.03 * smo + 0.06 * educ - 0.01 * hypertension + 0.05 * cardio | 0.58 | |
| hippocampus | -0.17 - 0.19 * age - 0.04 * apoe4 + 0.56 * sex + 0.09 * bmi - 0.27 * cogn + 0.15 * aβ + 0.34 * alcohol - 0.37 * tau | 0.49 | |
| ventricles | -0.22 + 0.34 * cogn - 0.14 * aβ - 0.52 * alcohol | 0.43 | |
| icv | -0.62 - 0.00 * age + 1.03 * sex + 0.17 * cogn - 0.03 * Aβ - 0.25 * tau | 0.61 | |
| mmse | 0.15 - 0.41 * cogn + 0.11 * education - 0.04 * age + 0.02 * Aβ + 0.03 * tau + 0.25 * fdg + 0.05 * ventricles + 0.18 * hippocampus + 0.05 * icv | 0.44 | |
| Additional identified correlations, which were not specified in the DAG. | | | |
| s | 0.06 * age | | |
| a | -0.05 * APOE ε4 | | |

Table S4: Summary statistics of variables in generated training and test datasets. Each dataset was simulated with n=10,000 observations using estimated parameters from structural equation modelling (SEM). SEM was applied on three imputed datasets. This table summarizes results generated using SEM-parameters from the first imputed dataset. Internal validation datasets were simulated by sampling using original distributions of the exogenous variables age, APOE ε4, and sex from ADNI and linear equations from SEM. Intervention test sets were generated by reducing the mean age, APOE ε4 frequency or changing the CSF-tau mechanism and applying linear equations with parameters estimated from SEM. The mean of each numeric variable was recorded for each simulation. The medians, 2.5 and 97.5-percentiles were calculated across means for 10,000 simulations and are being compared to the mean values in the original dataset.

| variable | original dataset | internal validation, median (2.5-97.5%) | age intervention means, median (2.5-97.5%) | age 2 intervention means, median (2.5-97.5%) | APOE ε4 intervention, median (2.5-97.5%) | tau intervention, median (2.5-97.5%) |
|---|---|---|---|---|---|---|
| age | 73.8 | 73.8 (73.6-73.9) | 35.0 (34.8-35.2) | 65.0 (64.8-65.2) | 73.8 (73.6-73.9) | 73.8 (73.6-73.9) |
| APOE ε4 | 46.9 | 47.0 (46.0-48.0) | 47.0 (46.0-48.0) | 47.0 (46.0-48.0) | 5.0 (4.6-5.4) | 47.0 (46.0-47.9) |
| sex | 55.1 | 55.1 (54.1-56.1) | 55.1 (54.1-56.1) | 55.1 (54.1-56.1) | 55.1 (54.1-56.1) | 55.1 (54.1-56.1) |
| cognitive impairment | 69.9 | 70.0 (69.1-71.0) | 68.3 (67.4-69.3) | 69.6 (68.7-70.6) | 63.1 (62.1-64.1) | 82.3 (81.3-83.1) |
| education | 15.9 | 16.1 (16.0-16.2) | 17.6 (17.5-17.7) | 16.4 (16.4-16.5) | 16.1 (16.0-16.2) | 16.1 (16.0-16.2) |
| bmi | 29.1 | 29.7 (29.6-29.8) | 33.8 (33.7-33.9) | 30.7 (30.6-30.8) | 29.7 (29.6-29.8) | 29.7 (29.6-29.8) |
| history of smoking (%) | 24.3 | 23.4 (22.3-24.8) | 34.1 (33.1-35.3) | 25.7 (24.6-27.1) | 23.4 (22.3-24.9) | 23.3 (22.3-24.9) |
| history of alcohol abuse (%) | 2.5 | 1.5 (1.2-1.7) | 46.5 (44.6-49.5) | 5.3 (4.8-6.0) | 1.5 (1.2-1.7) | 1.5 (1.2-1.7) |
| history of hypertension (%) | 34.1 | 35.0 (34.2-36.1) | 8.0 (7.4-8.7) | 27.2 (26.2-28.1) | 35.1 (34.2-36.1) | 35.1 (34.2-36.1) |
| history of cardiovascular events (%) | 67.8 | 65.1 (64.1-66.0) | 32.4 (31.4-33.3) | 57.6 (56.6-58.6) | 65.1 (64.1-66.0) | 65.1 (64.1-66.0) |
| fdg-PET | 1.2 | 1.2 (1.2-1.2) | 1.3 (1.3-1.3) | 1.2 (1.2-1.3) | 1.3 (1.3-1.3) | 1.1 (1.1-1.1) |
| CSF-Aβ | 1052.7 | 1076.6 (1064.4-1088.1) | 1521.0 (1509.5-1533.2) | 1176.9 (1165.3-1189.0) | 1291.2 (1280.2-1302.1) | 1076.6 (1064.4-1088.1) |
| CSF-tau | 287.0 | 292.7 (290.1-295.2) | 180.8 (178.2-183.4) | 267.3 (264.8-269.9) | 260.1 (257.5-262.6) | 476.4 (471.9-480.9) |
| Mini Mental State Exam (mmse) | 27.2 | 27.3 (27.2-27.3) | 29.6 (29.6-29.8) | 27.8 (27.8-27.9) | 27.8 (27.7-27.8) | 26.0 (26.0-26.1) |

Table S5: Calibration and discrimination of algorithms predicting cognitive impairment, measured by Integrated Calibration Index (ICI), Brier calibration component and area under the receiver operating characteristic curve (AUC). Cognitive impairment was predicted using logistic regression, lasso regression, random forest (rf) and generalized boosted regression (gbm). Trained models were tested in an internal validation dataset and external validation datasets with either reduced age, APOE ε4-frequency (APOE ε4) or modified CSF-tau mechanism. Models made predictions using either all variables, only causes (parents), only consequences (children) of cognitive impairment, or only exogenous variables. The table summarizes results generated using SEM-parameters from the first imputed dataset. Calibration and discrimination performance of all models were calculated with median, 2.5% percentile, 97.5% percentile across the 10,000 repetitions.

| Intervention | Model | Predictors | ICI median | ICI CI2.5 | ICI CI97.5 | Brier median | Brier CI2.5 | Brier CI97.5 | AUC median | AUC CI2.5 | AUC CI97.5 |
|---|---|---|---|---|---|---|---|---|---|---|---|
| internal | logistic | all | 0.009 | 0.004 | 0.016 | 0.000 | 0.000 | 0.000 | 0.75 | 0.73 | 0.76 |
| internal | logistic | parents | 0.009 | 0.004 | 0.016 | 0.000 | 0.000 | 0.000 | 0.64 | 0.63 | 0.65 |
| internal | logistic | children | 0.009 | 0.004 | 0.017 | 0.000 | 0.000 | 0.000 | 0.71 | 0.69 | 0.72 |
| internal | logistic | exogenous | 0.009 | 0.004 | 0.018 | 0.000 | 0.000 | 0.000 | 0.50 | 0.50 | 0.50 |
| internal | lasso | all | 0.009 | 0.004 | 0.016 | 0.000 | 0.000 | 0.000 | 0.75 | 0.73 | 0.76 |
| internal | lasso | parents | 0.009 | 0.004 | 0.016 | 0.000 | 0.000 | 0.000 | 0.64 | 0.62 | 0.65 |
| internal | lasso | children | 0.009 | 0.004 | 0.016 | 0.000 | 0.000 | 0.000 | 0.70 | 0.69 | 0.72 |
| internal | lasso | exogenous | 0.009 | 0.004 | 0.018 | 0.000 | 0.000 | 0.000 | 0.50 | 0.50 | 0.50 |
| internal | rf | all | 0.021 | 0.013 | 0.029 | 0.001 | 0.000 | 0.001 | 0.73 | 0.72 | 0.74 |
| internal | rf | parents | 0.035 | 0.024 | 0.046 | 0.001 | 0.001 | 0.002 | 0.62 | 0.6 | 0.63 |
| internal | rf | children | 0.017 | 0.009 | 0.026 | 0.000 | 0.000 | 0.001 | 0.70 | 0.69 | 0.71 |
| internal | rf | exogenous | 0.292 | 0.277 | 0.305 | 0.088 | 0.080 | 0.094 | 0.50 | 0.50 | 0.50 |
| internal | gbm | all | 0.036 | 0.027 | 0.045 | 0.002 | 0.001 | 0.003 | 0.71 | 0.70 | 0.73 |
| internal | gbm | parents | 0.029 | 0.019 | 0.039 | 0.001 | 0.000 | 0.002 | 0.60 | 0.59 | 0.62 |
| internal | gbm | children | 0.017 | 0.010 | 0.026 | 0.000 | 0.000 | 0.001 | 0.70 | 0.69 | 0.71 |

| | | | | | | | | | | | |
|---|---|---|---|---|---|---|---|---|---|---|---|
| internal | gbm | exogenous | 0.010 | 0.004 | 0.018 | 0.000 | 0.000 | 0.000 | 0.50 | 0.50 | 0.50 |
| age | logistic | all | 0.041 | 0.008 | 0.082 | 0.002 | 0.000 | 0.009 | 0.73 | 0.7 | 0.76 |
| age | logistic | parents | 0.017 | 0.005 | 0.053 | 0.000 | 0.000 | 0.003 | 0.65 | 0.62 | 0.67 |
| age | logistic | children | 0.309 | 0.287 | 0.331 | 0.108 | 0.093 | 0.124 | 0.69 | 0.67 | 0.7 |
| age | logistic | exogenous | 0.019 | 0.005 | 0.058 | 0.000 | 0.000 | 0.004 | 0.5 | 0.5 | 0.59 |
| age | lasso | all | 0.036 | 0.007 | 0.077 | 0.002 | 0.000 | 0.008 | 0.73 | 0.7 | 0.76 |
| age | lasso | parents | 0.018 | 0.005 | 0.056 | 0.000 | 0.000 | 0.003 | 0.65 | 0.62 | 0.67 |
| age | lasso | children | 0.306 | 0.283 | 0.327 | 0.106 | 0.091 | 0.121 | 0.69 | 0.68 | 0.7 |
| age | lasso | exogenous | 0.017 | 0.006 | 0.048 | 0.000 | 0.000 | 0.002 | 0.5 | 0.5 | 0.59 |
| age | rf | all | 0.213 | 0.174 | 0.249 | 0.052 | 0.036 | 0.069 | 0.72 | 0.7 | 0.74 |
| age | rf | parents | 0.122 | 0.077 | 0.165 | 0.015 | 0.006 | 0.028 | 0.66 | 0.63 | 0.68 |
| age | rf | children | 0.274 | 0.239 | 0.307 | 0.080 | 0.060 | 0.101 | 0.68 | 0.66 | 0.7 |
| age | rf | exogenous | 0.260 | 0.120 | 0.308 | 0.075 | 0.022 | 0.104 | 0.5 | 0.5 | 0.5 |
| age | gbm | all | 0.237 | 0.211 | 0.262 | 0.064 | 0.051 | 0.078 | 0.72 | 0.71 | 0.73 |
| age | gbm | parents | 0.140 | 0.113 | 0.161 | 0.020 | 0.014 | 0.027 | 0.68 | 0.67 | 0.69 |
| age | gbm | children | 0.253 | 0.230 | 0.279 | 0.072 | 0.060 | 0.087 | 0.7 | 0.69 | 0.72 |
| age | gbm | exogenous | 0.033 | 0.006 | 0.141 | 0.001 | 0.000 | 0.021 | 0.5 | 0.5 | 0.6 |
| age2 | logistic | all | 0.010 | 0.004 | 0.019 | 0.000 | 0.000 | 0.001 | 0.75 | 0.73 | 0.76 |
| age2 | logistic | parents | 0.010 | 0.004 | 0.019 | 0.000 | 0.000 | 0.000 | 0.64 | 0.63 | 0.65 |
| age2 | logistic | children | 0.067 | 0.055 | 0.079 | 0.006 | 0.004 | 0.008 | 0.73 | 0.72 | 0.74 |
| age2 | logistic | exogenous | 0.010 | 0.004 | 0.020 | 0.000 | 0.000 | 0.001 | 0.5 | 0.5 | 0.52 |

| | | | | | | | | | | | |
|---|---|---|---|---|---|---|---|---|---|---|---|
| age2 | lasso | all | 0.010 | 0.004 | 0.018 | 0.000 | 0.000 | 0.000 | 0.75 | 0.73 | 0.76 |
| age2 | lasso | parents | 0.009 | 0.004 | 0.019 | 0.000 | 0.000 | 0.000 | 0.64 | 0.63 | 0.65 |
| age2 | lasso | children | 0.066 | 0.054 | 0.078 | 0.006 | 0.004 | 0.008 | 0.73 | 0.72 | 0.74 |
| age2 | lasso | exogenous | 0.010 | 0.004 | 0.020 | 0.000 | 0.000 | 0.000 | 0.5 | 0.5 | 0.51 |
| age2 | rf | all | 0.049 | 0.039 | 0.060 | 0.003 | 0.002 | 0.005 | 0.74 | 0.73 | 0.76 |
| age2 | rf | parents | 0.019 | 0.010 | 0.031 | 0.000 | 0.000 | 0.001 | 0.63 | 0.61 | 0.64 |
| age2 | rf | children | 0.070 | 0.058 | 0.082 | 0.007 | 0.005 | 0.009 | 0.72 | 0.71 | 0.73 |
| age2 | rf | exogenous | 0.282 | 0.254 | 0.302 | 0.082 | 0.068 | 0.092 | 0.5 | 0.5 | 0.5 |
| age2 | gbm | all | 0.050 | 0.039 | 0.061 | 0.003 | 0.002 | 0.004 | 0.74 | 0.73 | 0.75 |
| age2 | gbm | parents | 0.033 | 0.023 | 0.043 | 0.001 | 0.001 | 0.002 | 0.63 | 0.61 | 0.64 |
| age2 | gbm | children | 0.058 | 0.046 | 0.070 | 0.004 | 0.002 | 0.006 | 0.73 | 0.71 | 0.74 |
| age2 | gbm | exogenous | 0.012 | 0.005 | 0.029 | 0.000 | 0.000 | 0.002 | 0.5 | 0.5 | 0.52 |
| APOE ε4 | logistic | all | 0.010 | 0.004 | 0.018 | 0.000 | 0.000 | 0.000 | 0.75 | 0.74 | 0.76 |
| APOE ε4 | logistic | parents | 0.010 | 0.004 | 0.019 | 0.000 | 0.000 | 0.000 | 0.65 | 0.64 | 0.66 |
| APOE ε4 | logistic | children | 0.018 | 0.009 | 0.029 | 0.000 | 0.000 | 0.001 | 0.71 | 0.7 | 0.72 |
| APOE ε4 | logistic | exogenous | 0.010 | 0.004 | 0.021 | 0.000 | 0.000 | 0.000 | 0.5 | 0.5 | 0.5 |
| APOE ε4 | lasso | all | 0.009 | 0.004 | 0.017 | 0.000 | 0.000 | 0.000 | 0.75 | 0.74 | 0.76 |
| APOE ε4 | lasso | parents | 0.009 | 0.004 | 0.018 | 0.000 | 0.000 | 0.000 | 0.65 | 0.64 | 0.66 |
| APOE ε4 | lasso | children | 0.018 | 0.008 | 0.029 | 0.000 | 0.000 | 0.001 | 0.71 | 0.7 | 0.72 |
| APOE ε4 | lasso | exogenous | 0.010 | 0.003 | 0.021 | 0.000 | 0.000 | 0.000 | 0.5 | 0.5 | 0.5 |
| APOE ε4 | rf | all | 0.024 | 0.015 | 0.033 | 0.001 | 0.000 | 0.001 | 0.74 | 0.73 | 0.75 |

| | | | | | | | | | | | |
|---|---|---|---|---|---|---|---|---|---|---|---|
| APOE ε4 | rf | parents | 0.037 | 0.025 | 0.050 | 0.002 | 0.001 | 0.003 | 0.63 | 0.62 | 0.64 |
| APOE ε4 | rf | children | 0.022 | 0.013 | 0.031 | 0.001 | 0.000 | 0.001 | 0.7 | 0.69 | 0.71 |
| APOE ε4 | rf | exogenous | 0.356 | 0.336 | 0.370 | 0.128 | 0.114 | 0.136 | 0.5 | 0.5 | 0.5 |
| APOE ε4 | gbm | all | 0.037 | 0.027 | 0.047 | 0.002 | 0.001 | 0.003 | 0.73 | 0.72 | 0.74 |
| APOE ε4 | gbm | parents | 0.029 | 0.018 | 0.040 | 0.001 | 0.000 | 0.002 | 0.62 | 0.61 | 0.63 |
| APOE ε4 | gbm | children | 0.026 | 0.015 | 0.037 | 0.001 | 0.000 | 0.002 | 0.71 | 0.7 | 0.72 |
| APOE ε4 | gbm | exogenous | 0.011 | 0.004 | 0.022 | 0.000 | 0.000 | 0.001 | 0.5 | 0.5 | 0.5 |
| tau | logistic | all | 0.023 | 0.017 | 0.030 | 0.001 | 0.000 | 0.002 | 0.74 | 0.73 | 0.76 |
| tau | logistic | parents | 0.009 | 0.005 | 0.015 | 0.000 | 0.000 | 0.000 | 0.65 | 0.64 | 0.66 |
| tau | logistic | children | 0.037 | 0.028 | 0.045 | 0.002 | 0.001 | 0.003 | 0.75 | 0.74 | 0.77 |
| tau | logistic | exogenous | 0.117 | 0.105 | 0.128 | 0.017 | 0.014 | 0.020 | 0.5 | 0.5 | 0.51 |
| tau | lasso | all | 0.022 | 0.015 | 0.029 | 0.001 | 0.000 | 0.002 | 0.74 | 0.73 | 0.76 |
| tau | lasso | parents | 0.009 | 0.005 | 0.015 | 0.000 | 0.000 | 0.000 | 0.65 | 0.64 | 0.66 |
| tau | lasso | children | 0.037 | 0.029 | 0.045 | 0.002 | 0.001 | 0.003 | 0.75 | 0.74 | 0.77 |
| tau | lasso | exogenous | 0.117 | 0.105 | 0.128 | 0.016 | 0.013 | 0.020 | 0.5 | 0.5 | 0.5 |
| tau | rf | all | 0.026 | 0.017 | 0.036 | 0.001 | 0.000 | 0.002 | 0.74 | 0.73 | 0.75 |
| tau | rf | parents | 0.016 | 0.006 | 0.029 | 0.000 | 0.000 | 0.001 | 0.63 | 0.62 | 0.65 |
| tau | rf | children | 0.043 | 0.034 | 0.053 | 0.003 | 0.002 | 0.004 | 0.75 | 0.73 | 0.76 |
| tau | rf | exogenous | 0.175 | 0.158 | 0.190 | 0.033 | 0.029 | 0.037 | 0.5 | 0.5 | 0.5 |
| tau | gbm | all | 0.035 | 0.028 | 0.044 | 0.001 | 0.001 | 0.002 | 0.73 | 0.72 | 0.75 |
| tau | gbm | parents | 0.055 | 0.044 | 0.066 | 0.003 | 0.002 | 0.005 | 0.62 | 0.61 | 0.64 |

| | | | | | | | | | | | |
|---|---|---|---|---|---|---|---|---|---|---|---|
| tau | gbm | children | 0.047 | 0.039 | 0.056 | 0.002 | 0.002 | 0.003 | 0.74 | 0.73 | 0.76 |
| tau | gbm | exogenous | 0.117 | 0.106 | 0.129 | 0.016 | 0.013 | 0.020 | 0.5 | 0.5 | 0.51 |

Table S6: Transportability of algorithms trained to predict cognitive impairment, measured by differences of Integrated Calibration Index (ICI), Brier calibration component, and area under the receiver operating characteristic curve (AUC) between internal validation and intervention test sets. Three types of algorithms (logistic regression, random forest (rf) and generalized boosted regression (GBM)) were trained to predict cognitive impairment. Trained algorithms were applied predict cognitive impairment on validation datasets, which had the same distribution of age, APOE ε4 and sex variables, as in the training data and on intervention test set, which had either a younger population mean age (age and age2), lower APOE ε4 allele frequency (APOE ε4) or different CSF-tau (tau) mechanism compared to the training and internal validation datasets. Training and test data was generated with n=10,000 observations, 10,000 times based on obtained SEM-parameters of the first imputed dataset. Transportability was measured by the ICI, Brier calibration component, and AUC difference between internal validation and intervention sets. ICI, Brier, and AUC differences are given as median, 2.5% percentile and 97.5% percentile across 10,000 repetitions. Negative differences in ICI and Brier, and positive differences in AUC, indicate reduced transportability of the model in the intervention setting compared to the internal validation setting.

| **Intervention** | **Model** | **Predictors** | **ICI median diff.** | **ICI CI2.5 diff.** | **ICI CI97.5 diff.** | **Brier median diff..** | **Brier CI2.5 diff.** | **Brier CI97.5 diff.** | **AUC median diff** | **AUC CI2.5 diff.** | **AUC CI97.5 diff.** |
|---|---|---|---|---|---|---|---|---|---|---|---|
| age | logistic | all | -0.031 | -0.073 | 0.002 | -0.002 | -0.009 | 0.000 | 0.02 | -0.01 | 0.05 |
| age | logistic | parents | -0.009 | -0.045 | 0.006 | 0.000 | -0.003 | 0.000 | -0.01 | -0.04 | 0.02 |
| age | logistic | children | -0.300 | -0.322 | -0.276 | -0.108 | -0.123 | -0.093 | 0.02 | 0.00 | 0.04 |
| age | logistic | exogenous | -0.009 | -0.048 | 0.007 | 0.000 | -0.004 | 0.000 | 0.00 | -0.09 | 0.00 |
| age | lasso | all | -0.027 | -0.069 | 0.002 | -0.002 | -0.008 | 0.000 | 0.01 | -0.02 | 0.05 |
| age | lasso | parents | -0.009 | -0.047 | 0.006 | 0.000 | -0.003 | 0.000 | -0.01 | -0.04 | 0.02 |
| age | lasso | children | -0.297 | -0.319 | -0.273 | -0.106 | -0.121 | -0.091 | 0.02 | 0.00 | 0.03 |
| age | lasso | exogenous | -0.008 | -0.038 | 0.007 | 0.000 | -0.002 | 0.000 | 0.00 | -0.09 | 0.00 |
| age | rf | all | -0.192 | -0.229 | -0.151 | -0.051 | -0.068 | -0.036 | 0.00 | -0.02 | 0.03 |
| age | rf | parents | -0.087 | -0.133 | -0.039 | -0.014 | -0.027 | -0.005 | -0.05 | -0.07 | -0.02 |
| age | rf | children | -0.257 | -0.289 | -0.222 | -0.079 | -0.101 | -0.060 | 0.01 | -0.01 | 0.04 |

| | | | | | | | | | | | |
|---|---|---|---|---|---|---|---|---|---|---|---|
| age | rf | exogenous | 0.031 | -0.017 | 0.172 | 0.013 | -0.016 | 0.066 | 0.00 | 0.00 | 0.00 |
| age | gbm | all | -0.201 | -0.229 | -0.171 | -0.063 | -0.077 | -0.049 | -0.01 | -0.03 | 0.02 |
| age | gbm | parents | -0.111 | -0.137 | -0.080 | -0.019 | -0.026 | -0.012 | -0.07 | -0.09 | -0.06 |
| age | gbm | children | -0.236 | -0.264 | -0.210 | -0.071 | -0.087 | -0.059 | -0.01 | -0.03 | 0.01 |
| age | gbm | exogenous | -0.023 | -0.131 | 0.005 | -0.001 | -0.021 | 0.000 | 0.00 | -0.10 | 0.00 |
| age2 | logistic | all | -0.001 | -0.010 | 0.007 | 0.000 | 0.000 | 0.000 | 0.00 | -0.02 | 0.01 |
| age2 | logistic | parents | -0.001 | -0.011 | 0.008 | 0.000 | 0.000 | 0.000 | 0.00 | -0.02 | 0.01 |
| age2 | logistic | children | -0.058 | -0.070 | -0.042 | -0.006 | -0.008 | -0.004 | -0.03 | -0.04 | -0.01 |
| age2 | logistic | exogenous | -0.001 | -0.012 | 0.009 | 0.000 | 0.000 | 0.000 | 0.00 | -0.02 | 0.00 |
| age2 | lasso | all | -0.001 | -0.010 | 0.007 | 0.000 | 0.000 | 0.000 | 0.00 | -0.02 | 0.01 |
| age2 | lasso | parents | -0.001 | -0.011 | 0.008 | 0.000 | 0.000 | 0.000 | 0.00 | -0.02 | 0.01 |
| age2 | lasso | children | -0.057 | -0.070 | -0.041 | -0.006 | -0.008 | -0.004 | -0.03 | -0.04 | -0.01 |
| age2 | lasso | exogenous | -0.001 | -0.011 | 0.009 | 0.000 | 0.000 | 0.000 | 0.00 | -0.01 | 0.00 |
| age2 | rf | all | -0.028 | -0.040 | -0.016 | -0.002 | -0.004 | -0.001 | -0.01 | -0.03 | 0.00 |
| age2 | rf | parents | 0.015 | 0.000 | 0.029 | 0.001 | 0.000 | 0.002 | -0.01 | -0.03 | 0.01 |
| age2 | rf | children | -0.053 | -0.065 | -0.040 | -0.006 | -0.009 | -0.004 | -0.02 | -0.04 | -0.01 |
| age2 | rf | exogenous | 0.010 | -0.012 | 0.039 | 0.006 | -0.005 | 0.019 | 0.00 | 0.00 | 0.00 |
| age2 | gbm | all | -0.014 | -0.029 | 0.001 | -0.001 | -0.003 | 0.001 | -0.02 | -0.04 | -0.01 |
| age2 | gbm | parents | -0.004 | -0.018 | 0.009 | 0.000 | -0.001 | 0.001 | -0.02 | -0.04 | -0.01 |
| age2 | gbm | children | -0.041 | -0.055 | -0.025 | -0.004 | -0.005 | -0.002 | -0.03 | -0.04 | -0.01 |
| age2 | gbm | exogenous | -0.002 | -0.020 | 0.008 | 0.000 | -0.002 | 0.000 | 0.00 | -0.02 | 0.00 |

| | | | | | | | | | | | |
|---|---|---|---|---|---|---|---|---|---|---|---|
| APOE ε4 | logistic | all | -0.001 | -0.009 | 0.007 | 0.000 | 0.000 | 0.000 | -0.01 | -0.02 | 0.01 |
| APOE ε4 | logistic | parents | -0.001 | -0.010 | 0.008 | 0.000 | 0.000 | 0.000 | -0.01 | -0.03 | 0.00 |
| APOE ε4 | logistic | children | -0.009 | -0.019 | 0.002 | 0.000 | -0.001 | 0.000 | -0.01 | -0.02 | 0.01 |
| APOE ε4 | logistic | exogenous | -0.001 | -0.012 | 0.008 | 0.000 | 0.000 | 0.000 | 0.00 | 0.00 | 0.00 |
| APOE ε4 | lasso | all | -0.001 | -0.009 | 0.007 | 0.000 | 0.000 | 0.000 | -0.01 | -0.02 | 0.00 |
| APOE ε4 | lasso | parents | 0.000 | -0.010 | 0.008 | 0.000 | 0.000 | 0.000 | -0.01 | -0.03 | 0.00 |
| APOE ε4 | lasso | children | -0.009 | -0.020 | 0.002 | 0.000 | -0.001 | 0.000 | -0.01 | -0.02 | 0.01 |
| APOE ε4 | lasso | exogenous | 0.000 | -0.011 | 0.009 | 0.000 | 0.000 | 0.000 | 0.00 | 0.00 | 0.00 |
| APOE ε4 | rf | all | -0.003 | -0.012 | 0.007 | 0.000 | -0.001 | 0.000 | -0.01 | -0.03 | 0.00 |
| APOE ε4 | rf | parents | -0.002 | -0.015 | 0.010 | 0.000 | -0.001 | 0.001 | -0.02 | -0.03 | 0.00 |
| APOE ε4 | rf | children | -0.005 | -0.017 | 0.007 | 0.000 | -0.001 | 0.000 | -0.01 | -0.02 | 0.01 |
| APOE ε4 | rf | exogenous | -0.063 | -0.080 | -0.046 | -0.039 | -0.049 | -0.029 | 0.00 | 0.00 | 0.00 |
| APOE ε4 | gbm | all | -0.001 | -0.011 | 0.008 | 0.000 | -0.001 | 0.001 | -0.01 | -0.03 | 0.00 |
| APOE ε4 | gbm | parents | 0.000 | -0.011 | 0.011 | 0.000 | -0.001 | 0.001 | -0.02 | -0.03 | 0.00 |
| APOE ε4 | gbm | children | -0.009 | -0.020 | 0.002 | 0.000 | -0.001 | 0.000 | -0.01 | -0.02 | 0.01 |
| APOE ε4 | gbm | exogenous | -0.001 | -0.012 | 0.009 | 0.000 | 0.000 | 0.000 | 0.00 | 0.00 | 0.00 |
| tau | logistic | all | -0.014 | -0.022 | -0.005 | -0.001 | -0.002 | 0.000 | 0.00 | -0.01 | 0.02 |
| tau | logistic | parents | 0.000 | -0.007 | 0.008 | 0.000 | 0.000 | 0.000 | -0.01 | -0.03 | 0.00 |
| tau | logistic | children | -0.028 | -0.037 | -0.015 | -0.002 | -0.003 | -0.001 | -0.05 | -0.06 | -0.03 |
| tau | logistic | exogenous | -0.108 | -0.120 | -0.092 | -0.017 | -0.020 | -0.014 | 0.00 | -0.01 | 0.00 |
| tau | lasso | all | -0.013 | -0.022 | -0.005 | -0.001 | -0.001 | 0.000 | 0.00 | -0.01 | 0.02 |

| | | | | | | | | | | | |
|---|---|---|---|---|---|---|---|---|---|---|---|
| tau | lasso | parents | 0.000 | -0.008 | 0.008 | 0.000 | 0.000 | 0.000 | -0.01 | -0.03 | 0.00 |
| tau | lasso | children | -0.028 | -0.038 | -0.016 | -0.002 | -0.003 | -0.001 | -0.05 | -0.06 | -0.03 |
| tau | lasso | exogenous | -0.108 | -0.120 | -0.092 | -0.016 | -0.019 | -0.012 | 0.00 | 0.00 | 0.00 |
| tau | rf | all | -0.005 | -0.017 | 0.006 | 0.000 | -0.001 | 0.000 | -0.01 | -0.03 | 0.01 |
| tau | rf | parents | 0.018 | -0.001 | 0.036 | 0.001 | 0.000 | 0.002 | -0.02 | -0.03 | 0.00 |
| tau | rf | children | -0.026 | -0.038 | -0.014 | -0.003 | -0.004 | -0.001 | -0.05 | -0.06 | -0.03 |
| tau | rf | exogenous | 0.117 | 0.099 | 0.134 | 0.054 | 0.048 | 0.061 | 0.00 | 0.00 | 0.00 |
| tau | gbm | all | 0.000 | -0.010 | 0.011 | 0.000 | 0.000 | 0.001 | -0.02 | -0.03 | 0.00 |
| tau | gbm | parents | -0.026 | -0.039 | -0.014 | -0.002 | -0.004 | -0.001 | -0.02 | -0.03 | 0.00 |
| tau | gbm | children | -0.031 | -0.041 | -0.018 | -0.002 | -0.003 | -0.001 | -0.04 | -0.06 | -0.03 |
| tau | gbm | exogenous | -0.108 | -0.120 | -0.092 | -0.016 | -0.020 | -0.013 | 0.00 | -0.01 | 0.00 |

### Supplementary Figures

Supplementary Figure S1: Initially created directed acyclic graph (DAG) that was created and tested using conditional independence tests. Predictor variables are marked in blue and the outcome variable (cognitive status) in green. Directed arrows indicate assumed causal relationships between variables. The included variables are: APOE ε4 (apoe4), age, sex, education (educ), CSF-Aβ (aβ), history of alcohol abuse (alc), history of smoking behavior (smok), Body Mass Index (bmi), history of hypertension (hypert), CSF-tau (tau), history of cardiovascular events (cardio), cognitive status (cogn), hippocampus (hippo), ventricles (ventr), intracranial volume (icv), FDG-PET (fdg), Mini-Mental State Exam score (mmse).

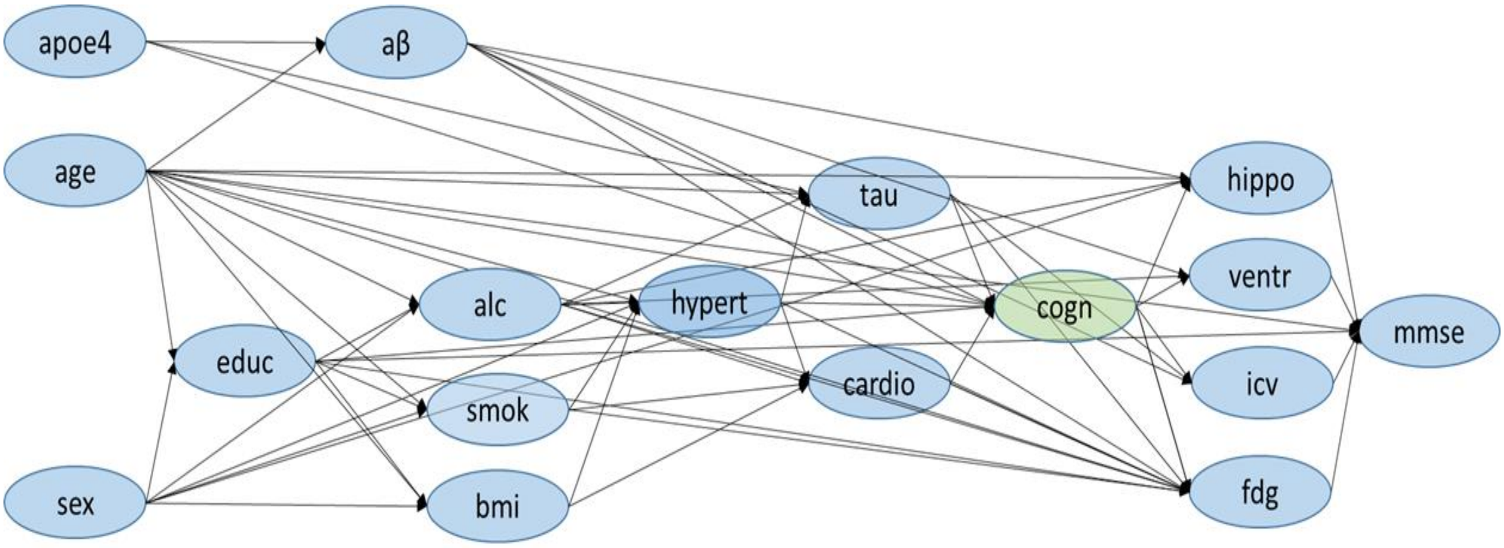

Supplementary Figure S2: Overview of process to generate datasets for training and validating prediction models. Each dataset consists of the exogenous (independent) variables age, sex and APOE ε4 and 14 endogenous variables. The exogenous variables served as input to generate endogenous variables. Endogenous variables were generated one by one by using the defined linear equations with obtained parameter estimates from the structural equation model (SEM) and the generated exogenous and endogenous variables. The exogenous variables for the training and internal validation dataset were sampled by bootstrapping the respective variable in the original ADNI dataset 10,000 times, to achieve that the distribution of the simulated data for training and internal validation matches the ADNI data. For the age-intervention settings (age and age2), age values were randomly sampled from a normal distribution with defined mean (age: 35, age2: 65) and std. deviation (age and age2: 10), while sex and APOE ε4 values were generated from bootstrapping from the ADNI dataset. For the APOE ε4 intervention setting, the binary APOE ε4 values were sampled from a Bernoulli distribution with probability 0.05, while age and sex values were bootstrapped from the ADNI data. For the tau intervention setting, all three exogenous variables were bootstrapped from the original data, but the parameter estimates for the linear equation with tau as dependent variable were altered to generate the tau values and consequences of tau.

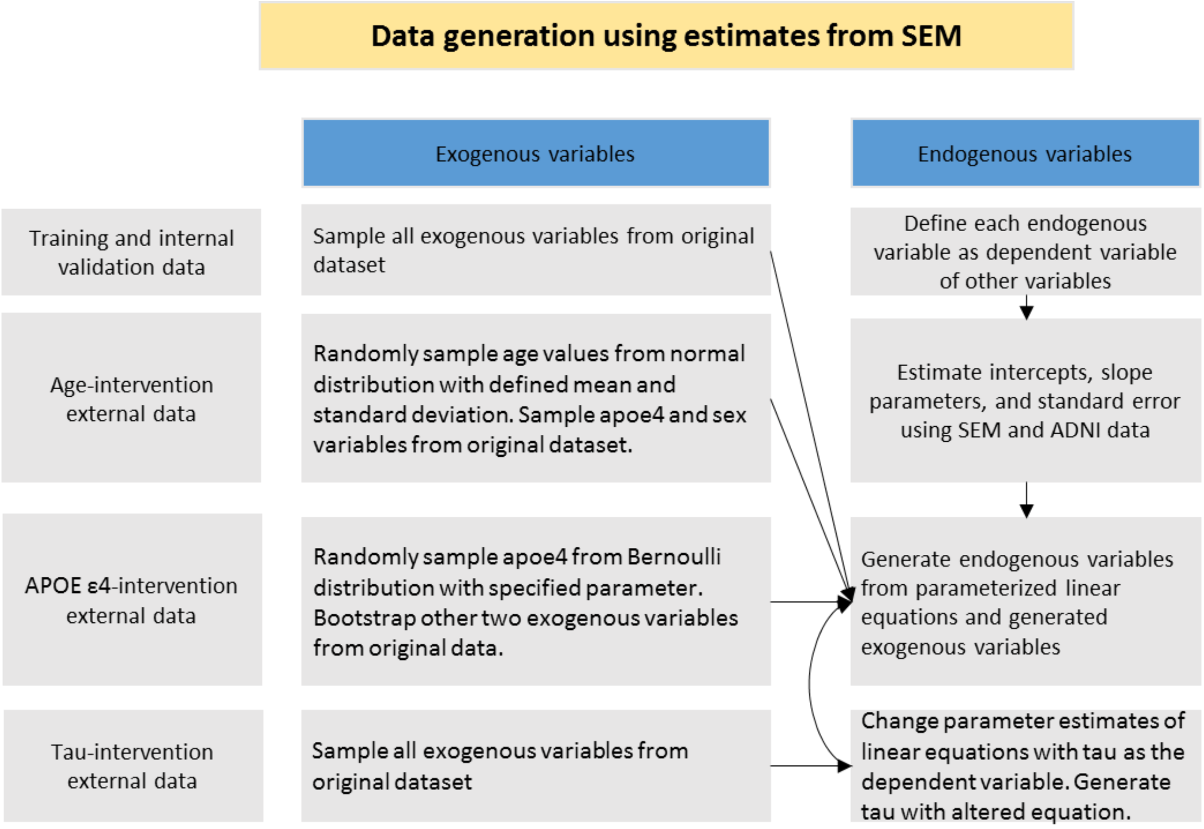

Supplementary Figure S3: Calibration in internal and external validation settings from 10,000 repetitions. The table summarizes results generated using SEM-parameters from the first imputed dataset. The figure shows calibration measured by the integrated calibration index (ICI) in the different external validation settings (age intervention, age2 intervention, APOE ε4 intervention, different CSF-tau mechanism). Cognitive impairment was predicted using logistic regression, lasso regression, random forest (rf) and generalized boosted regression (gbm) prediction models. Models were trained either with all predictor variables, only parent nodes (direct causes) of the outcome, only children nodes (consequences) of the outcome, or with the exogenous variables age, sex, and APOE ε4 (apoe4). Points in bottom left quadrant indicate models had good calibration in internal validation and external settings. Points in top left quadrant indicate models had poor calibration in external settings but good calibration in the internal validation. Points in the top right quadrant indicate poor calibration in both internal validation and external settings.

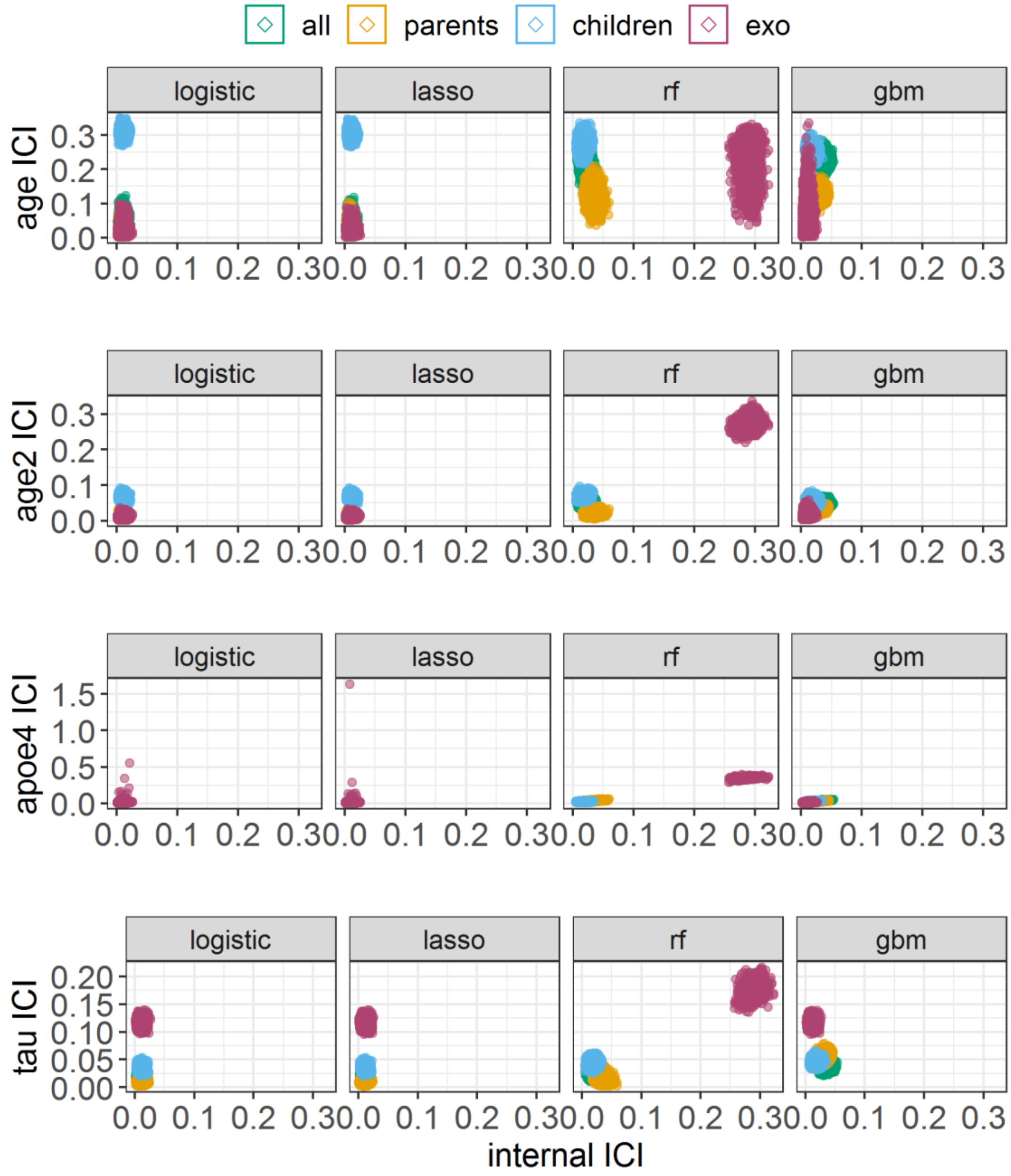

Supplementary Figure S4: Calibration in internal and external validation settings from 10,000 repetitions. The table summarizes results generated using SEM-parameters from the first imputed dataset. The figure shows calibration measured by the decomposed Brier score calibration component in the different external validation settings (age intervention, age2 intervention, APOE ε4 intervention, different CSF-tau mechanism Cognitive impairment was predicted using logistic regression, lasso regression, random forest (rf) and generalized boosted regression (gbm) prediction models. Models were trained either with all predictor variables, only parent nodes (direct causes) of the outcome, only children nodes (consequences) of the outcome, or with the exogenous variables age, sex, and APOE ε4 (apoe4). Points in bottom left quadrant indicate models had good calibration in internal validation and external settings. Points in top left quadrant indicate models had poor calibration in external settings but good calibration in the internal validation. Points in the top right quadrant indicate poor calibration in both internal validation and external settings.

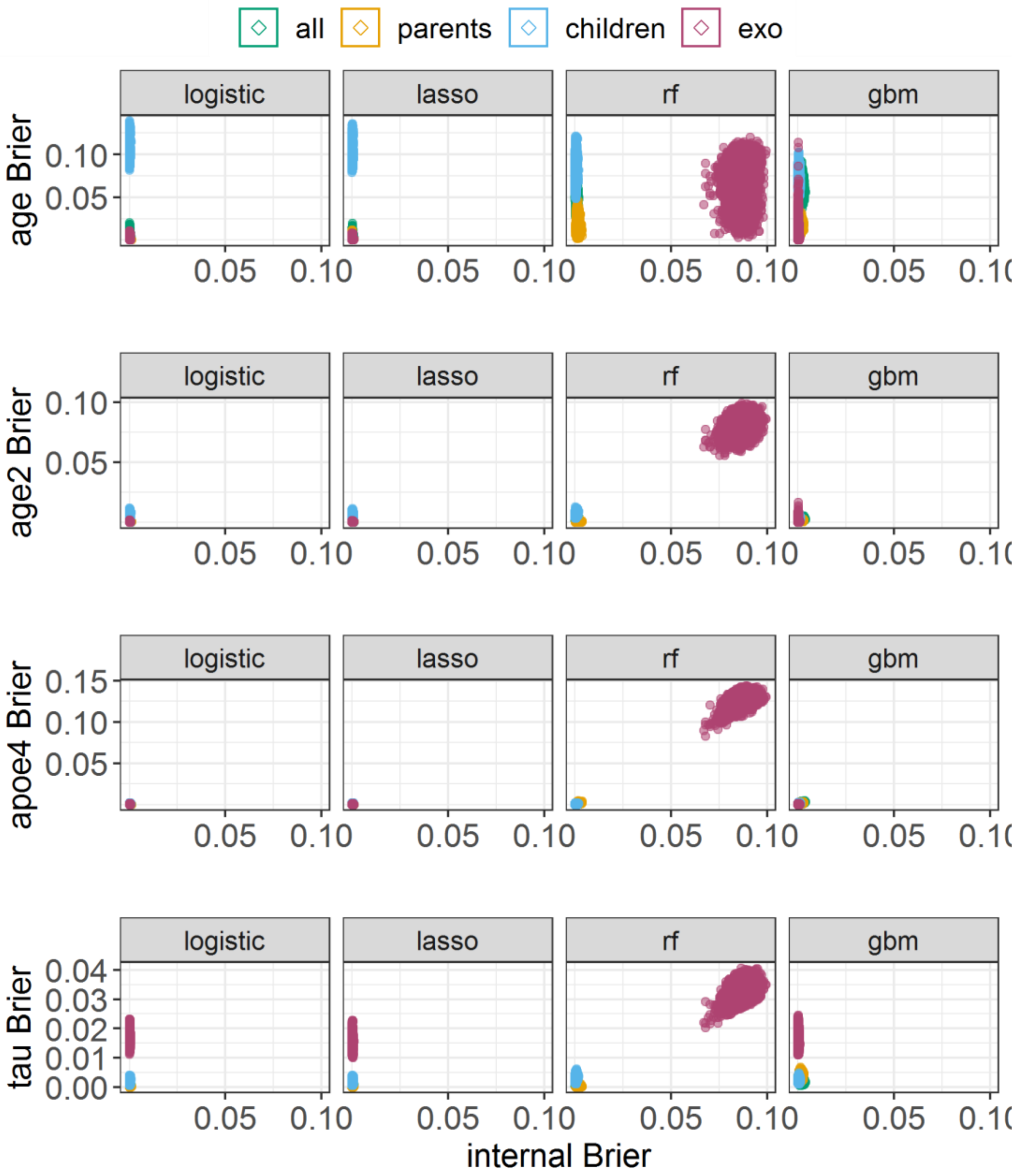

Supplementary Figure S5: Transportability between internal validation and external validation settings, measured by the difference of the Brier calibration component. The ADNI data to determine parameters for the generation of training and validation data, was imputed three times. Figure a) shows the results from 10,000 repetitions on the first imputed dataset, and Figure b) and c) from 10,000 repetitions on the other two imputed datasets.

**a)** **b)** **c)**

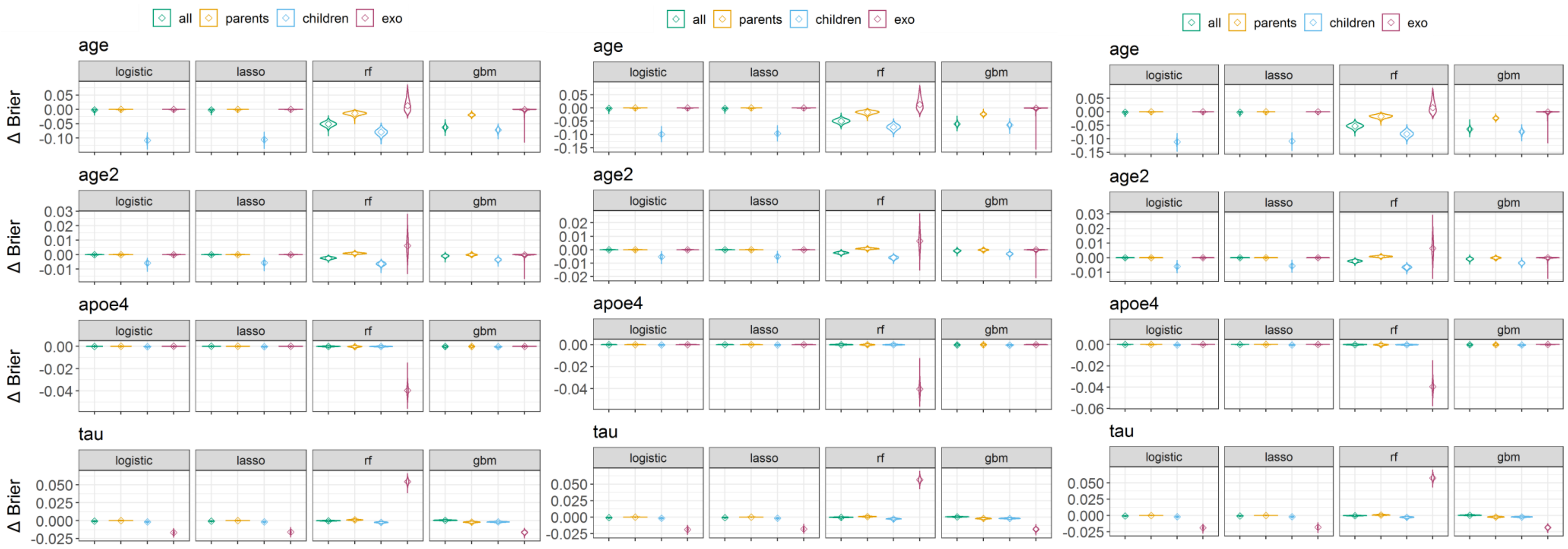

Supplementary Figure S6: Area under the receiver operating curve (AUC) in internal and external validation settings from 10,000 repetitions on one imputed dataset. Cognitive impairment was predicted using logistic regression, lasso regression, random forest (rf) and generalized boosted regression (gbm) prediction models. Models were trained either with all predictor variables, only parent nodes (direct causes) of the outcome, only children nodes (consequences) of the outcome, or with the exogenous variables age, sex, and APOE ε4 (apoe4). Points in bottom left quadrant indicate models had poor discrimination performance in internal validation and external settings. Points in top right quadrant indicate good discrimination performance in both internal validation and external settings.

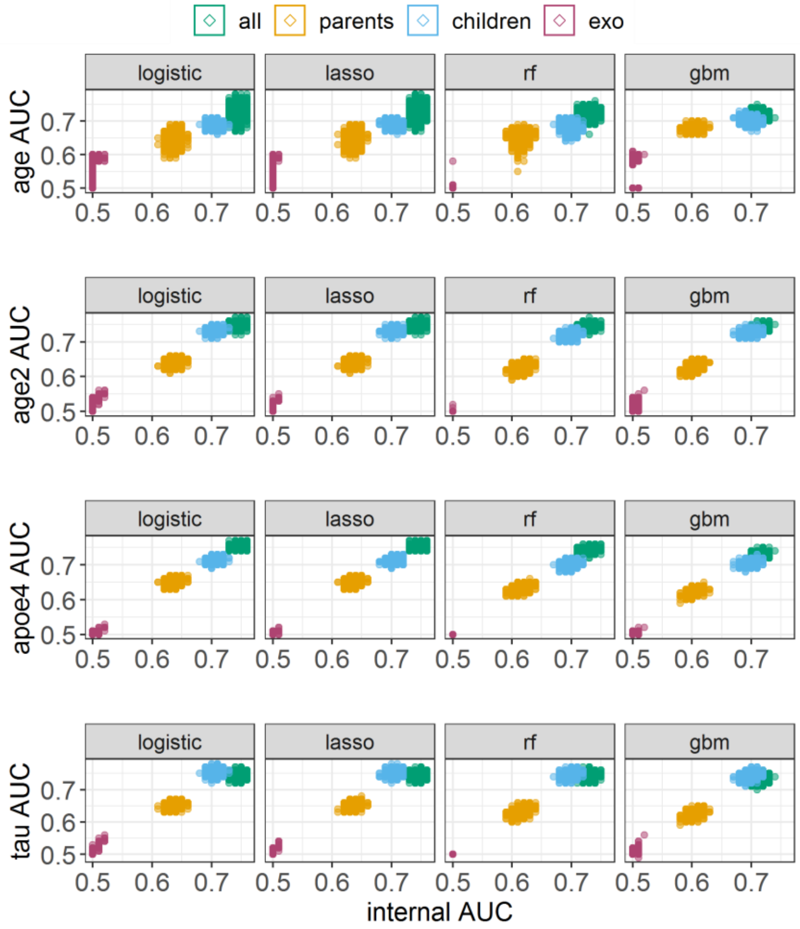

Supplementary Figure S7: Transportability between internal validation and external validation settings, measured by the difference of area under the receiver operating curve (AUC). The ADNI data to determine parameters for the generation of training and validation data, was imputed three times. Figure a) shows the results from 10,000 repetitions on the first imputed dataset, and Figure b) from 10,000 repetitions on another imputed dataset.

**a)**

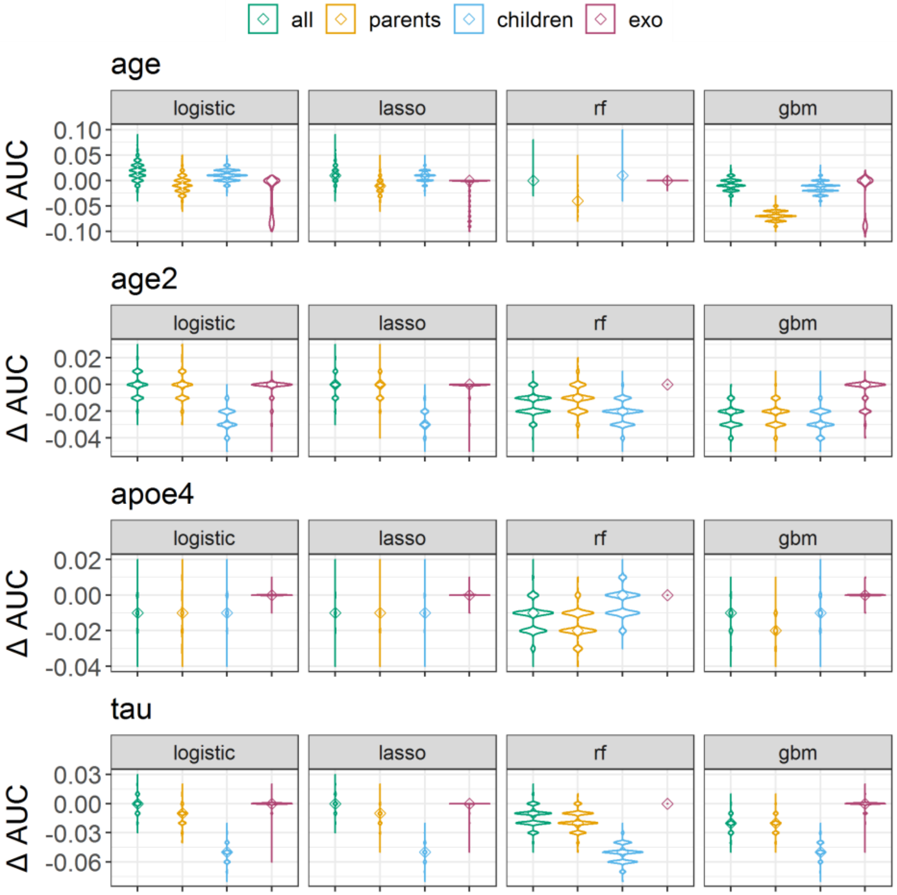

**b)**

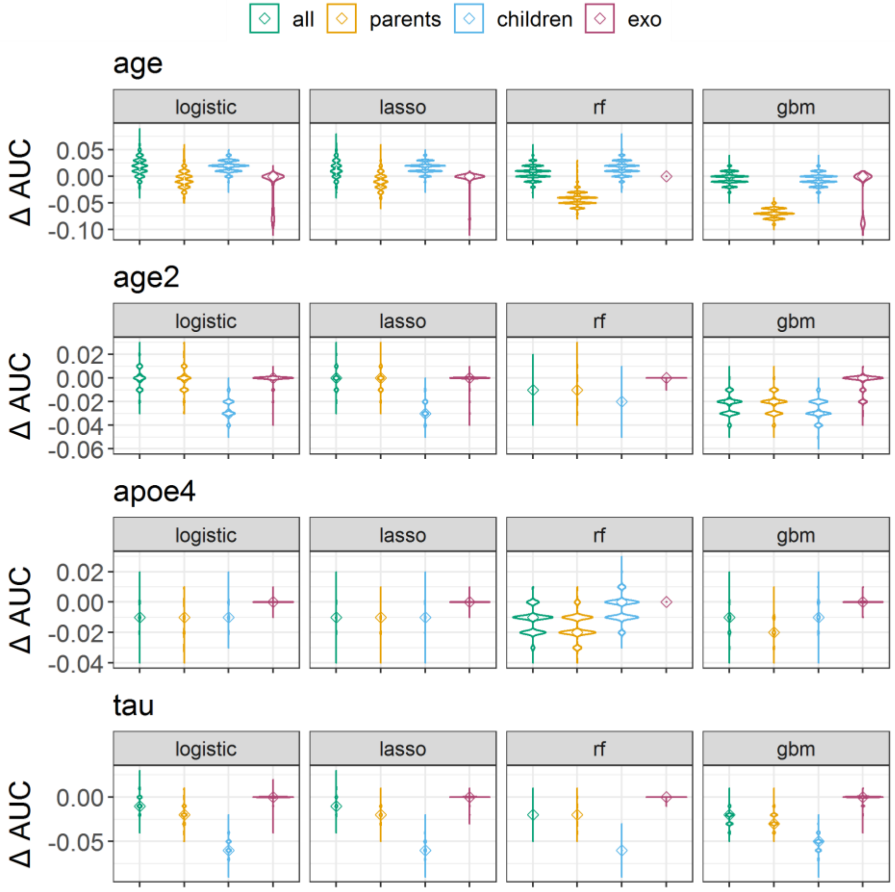

Supplementary Figure S8: Model performance in the internal validation setting, measured by the integrated calibration index (ICI), Brier calibration component, and area under the receiver-operating curve (AUC). The ADNI data to determine parameters for the generation of training and validation data, was imputed three times. Figure a) shows the results from 10,000 repetitions on the first imputed dataset, and Figure b) from 10,000 repetitions on another imputed dataset.

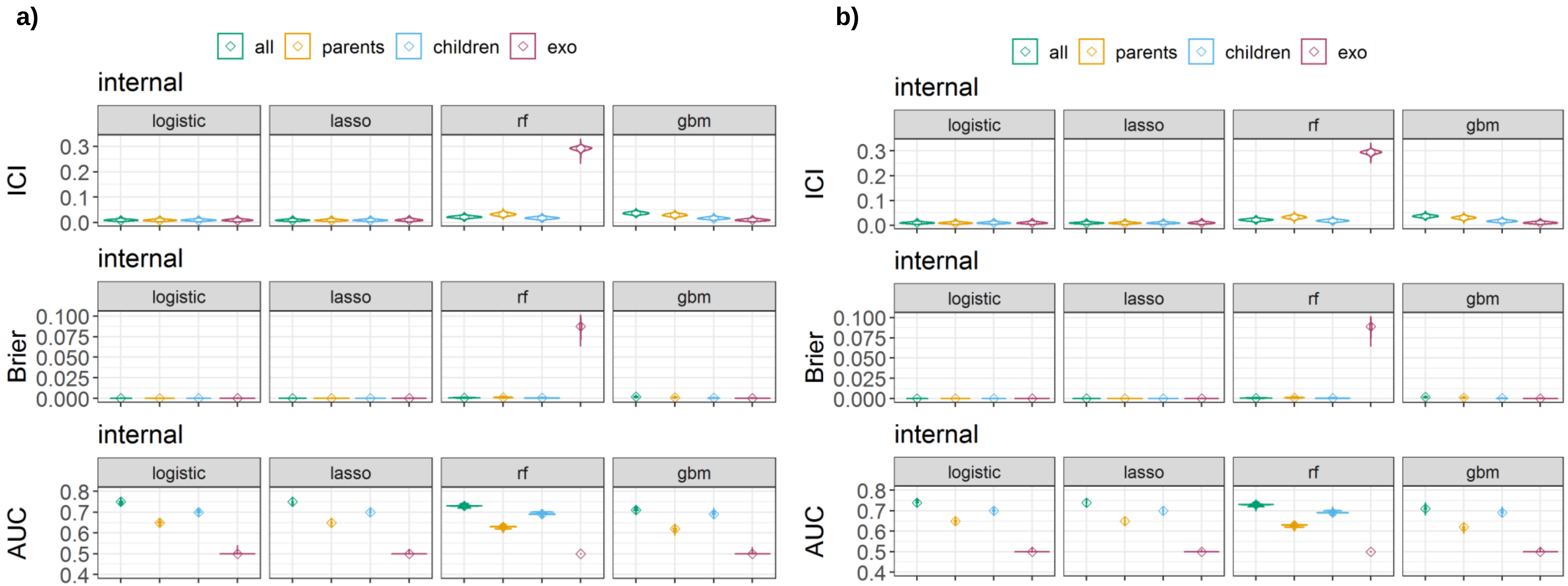

Supplementary Figure S9: Transportability between internal validation and external validation settings, measured by the difference of integrated calibration index (ICI). The ADNI data to determine parameters for the generation of training and validation data, was imputed three times. Figure a) shows the results from 10,000 repetitions on the first imputed dataset, and Figure b) from 10,000 repetitions on another imputed dataset.

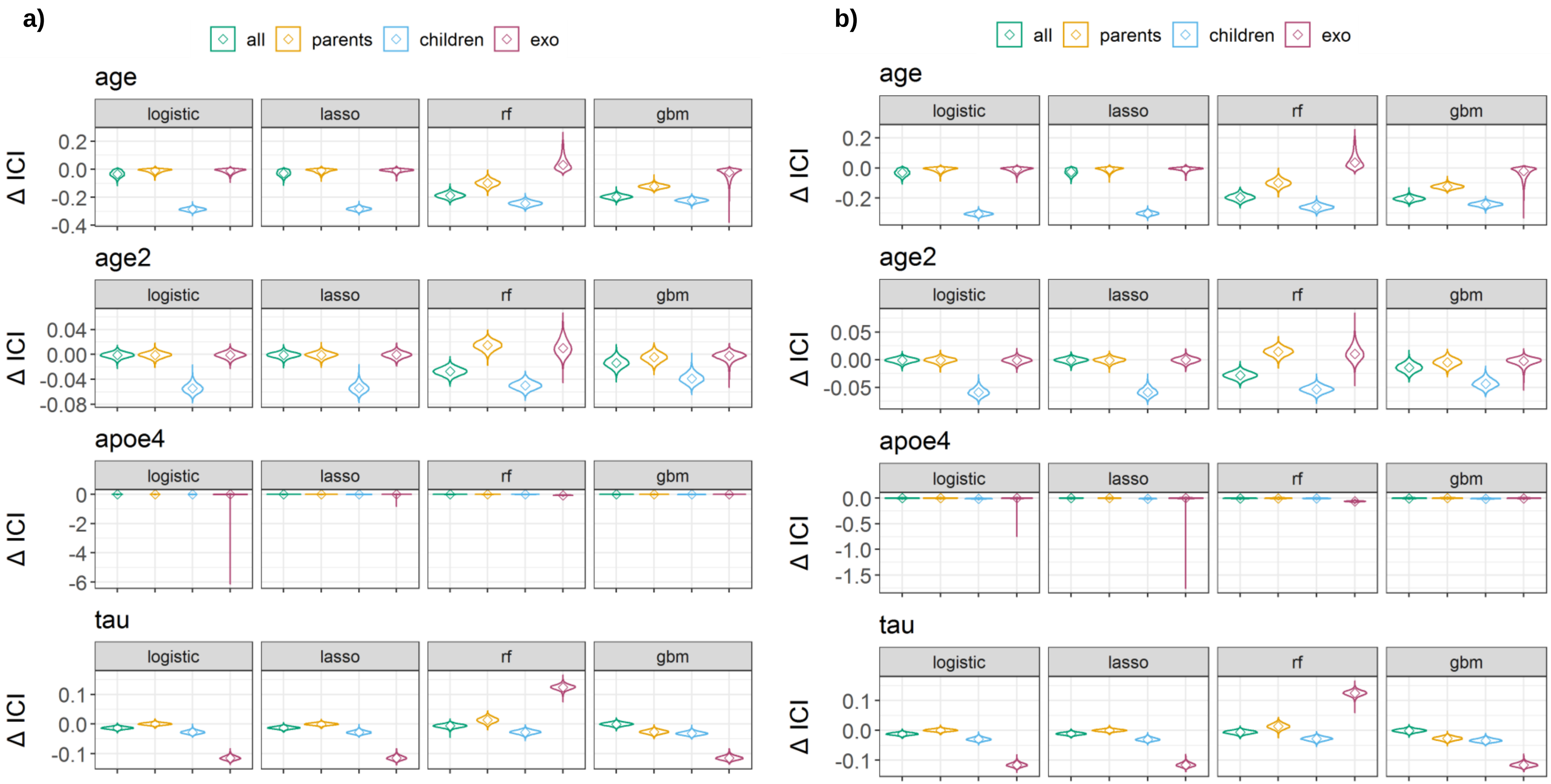

Supplementary Figure S10: Transportability of random forest (rf) models with optimized hyperparameter between internal validation and intervention test sets, measured by integrated calibration index (ICI), Brier calibration component and area under the receiver operating curve (AUC). Figure a) shows the performance in the internal validation setting. Figure b) shows the transportability, measured by differences between internal and external validation settings. The data for training and validating the random forest models was generated 100 times and hyperparameter were optimized for each training set to minimize the deviance. The optimized hyperparameter for the rf were number of predictors sampled for splitting at each node (mtry) from 1 to 5, and the minimum size of terminal nodes (nodesize) of 1, 5 or 10.

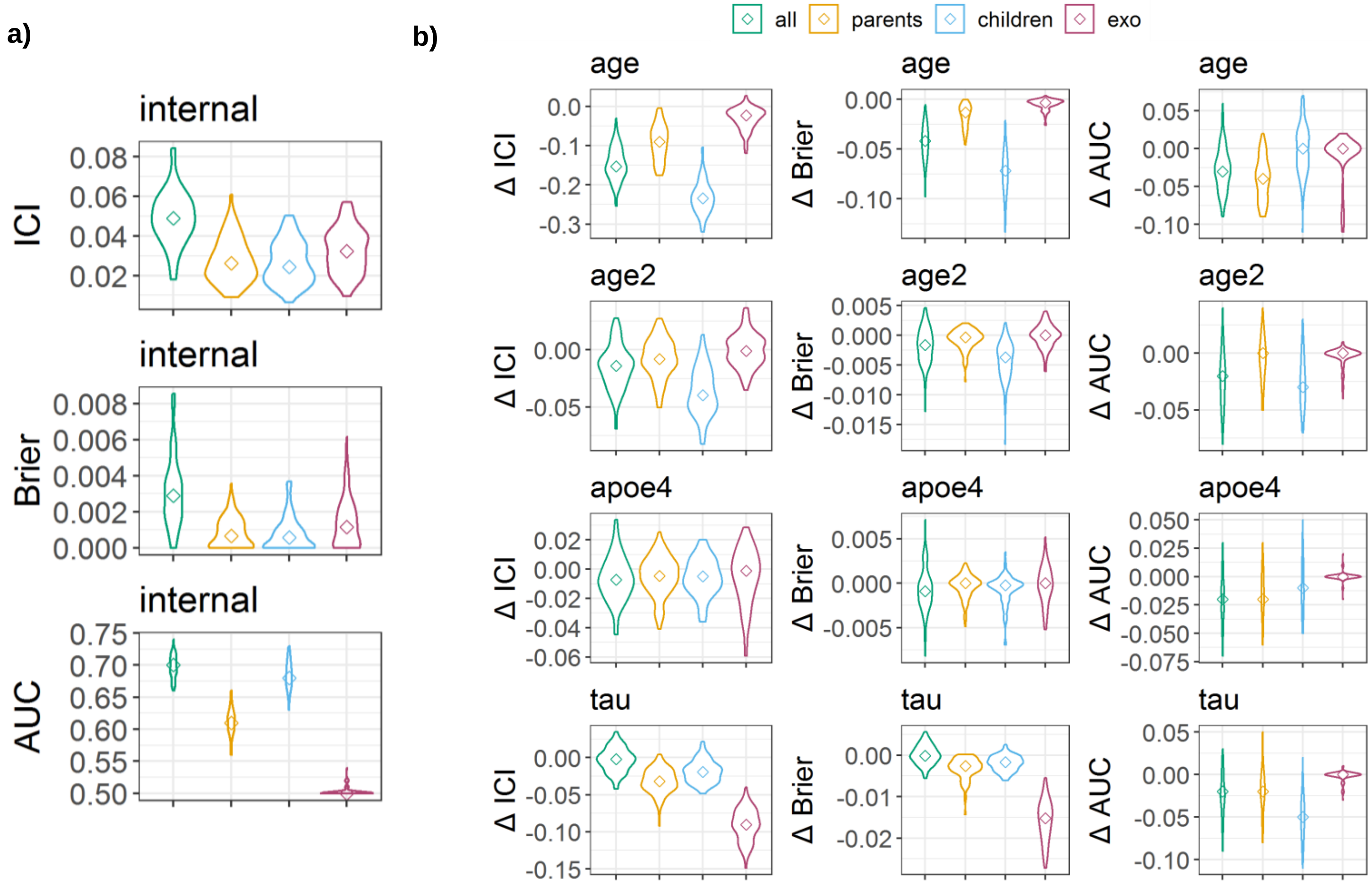
